# Soil surveillance for monitoring soil-transmitted helminth infections: method development and field testing in three countries

**DOI:** 10.1101/2023.09.26.23296174

**Authors:** Malathi Manuel, Heather K. Amato, Nils Pilotte, Benard Chieng, Sylvie B. Araka, Joël Edoux Eric Siko, Michael Harris, Maya Nadimpalli, Venkateshprabhu Janagaraj, Parfait Houngbegnon, Rajeshkumar Rajendiran, Joel Thamburaj, Saravanakumar Puthupalayam Kaliappan, Allison R. Sirois, Gretchen Walch, William E. Oswald, Kristjana H. Asbjornsdottir, Sean R. Galagan, Judd L. Walson, Steven A. Williams, Adrian J. F. Luty, Sammy M. Njenga, Moudachirou Ibikounlé, Sitara S.R. Ajjampur, Amy J. Pickering

## Abstract

One-fifth of the global population is infected with soil-transmitted helminths (STH). Mass drug administration (MDA) with deworming medication is widely implemented to control morbidity associated with STH infections. However, surveillance of human infection prevalence by collecting individual stool samples is time-consuming, costly, often stigmatized, and logistically challenging. Current methods of STH detection are poorly sensitive, particularly in low-intensity and low-prevalence populations. Here, we developed a sensitive and specific molecular method for detecting STH DNA in large volumes of soil by conducting laboratory and proof of concept studies across field sites in Kenya, Benin, and India. We collected human stool (n=669) and soil (n= 478) from 322 households across the three study sites. The overall prevalence of STH in soil was 31% for *Ascaris lumbricoides*, 3% for *T. trichuris*, and 24% for any hookworm species. Detection of an STH species in household soil was strongly associated with increased odds of a household member being infected with that species. Soil surveillance for STH has several benefits over stool-based surveillance, including lower cost and higher success rates for sample collection. Considering that delivery of MDA occurs at the community level, environmental surveillance using molecular methods could be a cost-effective alternate strategy for monitoring STH in these populations.

**Synopsis:** Limited data exists on the prevalence and reliability of environmental soil-transmitted helminth (STH) DNA as a marker of human infections in endemic populations. We developed a new molecular detection method for STH DNA in large-volume soil samples and field-tested it across three countries.

## 1. Introduction

Soil-transmitted helminths (STH) are a group of intestinal nematodes that include *Ascaris lumbricoides*, *Trichuris trichiura*, and the hookworm species, *Necator americanus* and *Ancylostoma duodenale*. STH infections are one of the most common infections among humans, affecting over 1.5 billion individuals globally, with children and pregnant women at highest risk for associated morbidity.^1^ STH are often endemic in low-income countries of Asia and Africa, where centralized or improved sanitation infrastructure remains limited in access.^1^ Infection occurs through ingestion of eggs of *A. lumbricoides* and *T. trichiura* (and occasionally *A. duodenale)* or larval penetration of the skin by hookworm larvae present in contaminated soil.^2^

The primary treatment strategy to date for controlling morbidity associated with STH infections in endemic settings is targeted mass drug administration (MDA).^3^ Recent evidence indicates that community-wide mass drug administration (cMDA) of all age groups with both high coverage and high adherence could potentially eliminate soil-transmitted helminth (STH) transmission in some settings.^4,5^ However, it remains unclear if STH transmission globally can be eliminated with cMDA strategy alone, or if MDA programs must be combined with improved sanitation programs and other forms of infrastructure and economic development.^6–8^

In settings with low coverage of networked sanitation and water supply infrastructure, persistent environmental reservoirs of STH eggs likely limit the effectiveness of MDA programs through increased chances for reinfection.^8–11^ A meta-analysis of studies from settings with medium-to-high endemic STH prevalence identified an average 12-month reinfection rate for *A. lumbricoides, T. trichiura* and hookworm of 94%, 82%, and 57%, respectively.^6^ Limited data on the prevalence and variability of STH in environmental reservoirs challenge our ability to account for environmental exposures to STH in MDA modeling estimations.^7,12^ Measurement of STH eggs in environmental soil within communities receiving cMDA would provide valuable additional data to model the effectiveness of deworming programs and community-level environmental characteristics which influence effectiveness.^12^ While STH control programs continue to rely on surveillance of human stool to assess STH prevalence within communities, sampling stool from individuals is resource intensive and logistically difficult to conduct. Developing specific and sensitive assays for detecting STH DNA in soil is a critical first step towards exploring if soil sampling could be a more cost-effective approach for monitoring the effectiveness of cMDA programs. Improved STH surveillance in the context of cMDA programs can help target programs to geographic areas where they are needed most, inform when cMDA is no longer needed, and trigger additional cMDA given early warning signs of recrudescence in the environment.

In recent years, quantitative polymerase chain reaction (qPCR) methods have achieved improved sensitivity for detecting STH infection as compared to Kato-Katz microscopy in human stool samples.^13,14^ In rural Kenya, considering all helminth species, qPCR was found to be more sensitive than Kato-Katz microscopy for STH identification in human stool samples.^15^ Additionally, qPCR assays can be designed to be species specific and can therefore exclude STH that infect animal hosts but are morphologically similar by microscopy.^16,17^

Here we optimize an STH DNA extraction protocol for large-quantity soil samples (20 g) and compare the performance of STH assays using qPCR and droplet digital PCR (ddPCR). We also compare our method to a previously published soil microscopy protocol to detect STH eggs^18^ and field-test our method on soil samples collected from household entrances and household drinking water sources in Benin, Kenya, and India. A comparison of soil STH results to human STH prevalence using stool samples collected from the same households was also performed. The results of this study will indicate whether molecular methods offer potential benefits for the monitoring of STH in the environment in endemic settings.

## 2. Materials & Methods

### 2.1 Study sites

This study was carried out at three sites: a rural commune in Comé, Benin, the rural health block of Timiri in Ranipet district of Tamil Nadu, India, and urban sub-counties of Kibra and Dagoretti South in Nairobi, Kenya. In each site, we aimed to enroll approximately 100 households for soil and human stool collection. The Benin and India sites were control clusters enrolled in an ongoing cluster randomized trial testing the feasibility of interrupting transmission of STH through expanded cMDA coverage (DeWorm3).^19,20^ The DeWorm3 study sites in Benin and India were previously censused, GIS mapped and divided into clusters, which were randomly assigned to control and treatment arms. Control arms received standard-of-care, school-based deworming (annual in Benin and bi-annual in India) and the intervention arms received bi-annual community-wide deworming with door-to-door drug distribution. This study leveraged the longitudinal monitoring cohort (LMC) in DeWorm3 which consisted of approximately 150 individuals per cluster from whom stool samples were collected annually. Among households with at least two LMC participants in control clusters, approximately 100 households were selected each in India and Benin during this study’s second year of sample collection. In Kenya, sample collection also supported a separate study investigating transmission of *Escherichia coli* across humans, poultry, and the environment.^21^ Eligible households had at least one child under 5 years old. The field team systematically approached households for inclusion in the study starting at a compound on a street known to have poultry in the vicinity; only one household was enrolled per compound. After enrolling a household, the field team walked to the next available compound and screened households as needed to ensure an equal number of households with and without poultry were enrolled on each street.

### 2.2 Survey and sample collection

Written informed consent was obtained from the head of the household or other adult with the ability to make decisions representing the household. IRB approval was obtained from Christian Medical College, Vellore (IRB Min no. 10392 dated 08.01.2018), the Ministry of Health in Benin (No. 15/MS/Dc/SGM/DRFMT/CNERS/SA), KEMRI Scientific and Ethics Review Unit (Protocol No. 3823), and Tufts Health Sciences Institutional Review Board (#13205). Household level surveys carried out at the time of sample collection captured data on socio-economic status (SES), access to safe water and sanitation, education, presence of animals, and deworming status. All data collection was carried out electronically with Android phones or Samsung tablets using SurveyCTO software (Dobility Inc.). In India and Benin, human stool samples were collected from individuals randomly selected into the LMC (the number of individuals ranged from one to four per household). In Kenya, up to three human stool samples were collected from the following age groups: 0-4, 5-14 and 15+ years. If it was not possible to collect a stool sample from each age group, the team collected either one additional stool sample from children 0-4 years of age or one additional stool sample from children 5-14 years of age. Households were visited up to three times to collect human stool samples. Soil samples were collected immediately outside the household entrance (within 2 meters). Soil samples were also collected within 2 m of the household’s reported primary drinking water source; only one sample was collected if multiple enrolled study households used the same water source. Using a stencil to mark off an area of 25 cm x 50 cm, approximately 100 g of soil was collected with a sterile scoop by scraping the top layer of the dirt inside the sampling area moving once vertically and then once horizontally. Both soil and human stool samples reached the laboratory within 3-4 hours of collection and were transported on ice packs.

### 2.3 Sample processing & physical soil characteristics

Once soil samples reached the laboratory, they were sieved through a screen to remove larger particles and then divided into three aliquots: 1) 20 g for DNA extraction (stored at -80°C); 2) 30 g for soil type, pH, and moisture content measurement (stored at 4°C); and 3) 15 g for soil microscopy (stored at 4°C). Soil pH was measured with a portable pH meter (Fisherbrand™ accumet™ AP110) after mixing 5 g of soil with 5 ml of distilled water and incubating at room temperature overnight. Soil moisture content was measured by weighing a soil sample, with an initial mass of 25 g, prior to and after placement in a hot air oven at 110°C for 16 hours. Soil type was categorized based on the ability to form a ball, ribbon and length of the ribbon formed using the same oven-dried soil sample mixed with a small amount of water.^22^

Human stool samples were mixed well and then aliquoted by weighing out 500mg of feces and placing it into a 2 mL cryovial containing 1 mL of 100% ethanol, followed by vortexing to homogenize. Stool samples were stored at -80°C until DNA extraction.

### 2.4 Soil microscopy

The soil samples were subjected to a series of filtration and flotation steps to concentrate any ova present using a previously published protocol.^18^ A 50 ml centrifuge tube containing 15 g of a sieved soil sample was filled to the 40 mL mark with 1% 7X solution (MP Biomedicals). The sample was then mixed well and incubated at room temperature overnight. Following incubation, each sample was vortexed and sieved (50 mesh, 300 µm, H&C sieving systems). The sieve was rinsed with 1% 7X solution and approximately 150 ml of the collected flow-through was allowed to settle at room temperature for 30 minutes. The supernatant was removed and the sediment was evenly divided into two 50 ml centrifuge tubes. The volume in each tube was then increased to 40 ml with 1% 7X solution and samples were centrifuged at 1000 x g for 10 minutes. Following centrifugation the supernatant was discarded. Five ml of zinc sulfate solution (1.25 specific gravity, flotation solution) was then added to each tube and samples were vortexed and centrifuged at 1000 x g for 5 minutes. The supernatant collected from both tubes was then combined and sieved a second time (500-mesh sieve, 25 µm, H&C sieving systems) and the contents recovered from the sieve were washed into a 50 ml tube with distilled water. The zinc sulfate flotation step was then repeated. Following this second flotation, the recovered solution was centrifuged at 1000 x g for 5 minutes and the supernatant was aspirated, leaving a 1 ml volume at the bottom of the tube. This entire 1 ml concentrate was then transferred to a Sedgewick Rafter slide (SPI supplies) and screened at 10X magnification. The morphology of the eggs identified was recorded, photographed and the number of eggs counted. If any STH eggs were putatively identified by microscopy, then the contents of the slide were washed back into a centrifuge tube using 1 ml of distilled water. Four ml of 0.1N sulphuric acid was then added to the tube and the sample was incubated at room temperature for 28 days. After 28 days the solution was centrifuged at 1000 x g for 2 minutes and the supernatant was aspirated, leaving 1 ml of the solution at the bottom of the tube. This residual volume was then screened for larvae to determine the viability of the eggs.

### 2.5 Molecular Analyses

#### Soil DNA extraction

Our goal was to develop a method that would enable processing a large quantity of soil to increase the chance of detecting DNA from STH eggs that can be present at concentrations <1 egg per gram of soil. The Qiagen DNeasy PowerMax Soil Kit was chosen based on the recommended input quantity of up to 10 g of soil. To further increase sensitivity, we modified the protocol to accommodate an initial homogenization and lysis step with 20 g of soil. Briefly, following the addition of soil samples to tubes containing PowerBead solution, the duration of homogenization was increased to 30 min on a vortexing platform; half of this solution was then processed (and the remaining half was discarded). An additional modification included re-loading and repeat centrifugation of extraction products following their final elution from Maxi Spin Columns. This post-elution re-exposure to the column was intended to maximize product recovery. We also modified the manufacturer’s protocol to include the addition of 100 pg of a previously described internal amplification control (IAC) plasmid.^23^ This plasmid was added to each sample following the addition of Solution C4.

Following isolation, samples underwent ethanol precipitation to further purify and concentrate the recovered DNA. To do so, 5 µL of Pellet Paint NF Co-precipitant (MilliporeSigma, Burlington, MA), 500 µL of 3M sodium acetate, and 10 mL of cold 100% ethanol were added to each elution product. Samples were vortexed briefly, incubated at room temperature for 2 min, and then centrifuged at maximum speed with the following conditions; 7,197 rcf for 5 mins in India, 8,500 rcf for 5 mins in Benin and 4,472 rcf for 10 mins in Kenya. Supernatant was then decanted, and 10 mL of cold 70% ethanol was added to each sample. Samples were again vortexed and centrifuged at maximum speed for 5 min. A second wash, this time using cold 100% ethanol, was then performed in an identical fashion. Following the aspiration of ethanol, pellets were allowed to air dry overnight, followed by resuspension in 200 µL of nuclease-free water.

All DNA extractions occurred in the countries in which the samples were collected. A reagent-only “extraction blank” sample was extracted after every 24 soil samples processed. The full extraction protocol is provided in Supporting Information (SI) (C).

#### Establishing limits of detection

Limits of detection (LOD) were established at Smith College using locally obtained, non-sample soil. To determine LODs for the qPCR-based detection of STH eggs in soil samples, 20 g aliquots of soil were spiked with either 200, 100, 50, 20 10, 5, or 2 *A. lumbricoides*, *N. americanus*, or *T. trichiura* eggs. Eggs were titrated from liquid suspensions with known concentrations. Following the addition of eggs to each sample, the full 20 g mass of each sample was thoroughly homogenized by hand mixing. Samples were processed in triplicate to determine the lowest spiking concentration that could be detected. DNA was extracted using the protocol described above for limit of detection testing.

#### Stool DNA extraction

DNA was extracted from stool samples using the MP Biomedicals FastDNA SPIN Kit for soil (MP Biomedicals) and a FastPrep benchtop homogenizer (MP Biomedicals) following a modified version^24^ of a previously published protocol.^25^ The same internal amplification control described in the soil sample extractions above was spiked in after lysis (100 pg IAC).^23^

#### Multi-parallel qPCR

Multi-parallel qPCR assays targeting non-coding repetitive sequences were utilized to detect *Necator americanus*, *Ancylostoma duodenale*, *Trichuris trichiura*,^14^ and *Ascaris lumbricoides^26^* in both soil and stool samples (SI, Table S8). Samples from India were additionally tested for the presence of *Ancylostoma ceylanicum*,^16^ a zoonotic species of hookworm known to contribute to human infection in many parts of Asia.^27^ All samples were tested in duplicate and a titration of plasmid (10 pg, 100 fg, and 1 fg) containing a single copy of the target sequence for each assay was utilized as a positive PCR control. ‘No template control’ samples were also tested on each qPCR reaction plate. For all the STH assays, cycling conditions included an initial 2 min incubation step at 50°C, followed by a 10 min incubation at 95°C, then 40 cycles of 15 sec at 95°C for denaturation and 1 min at 59°C for annealing and extension. The detailed protocol can be found in Supplemental Information (qPCR Protocol). All qPCR reactions were carried out using the Quantstudio 7 Flex PCR system (Applied Biosystems) and the data generated was analyzed using Quantstudio^TM^ Real-Time PCR software Version 1.3. A sample with a Cq value <40 in both replicates was reported as positive for the target tested. A sample returning a positive result in only one of two test replicates was re-tested, again in duplicate, and was reported positive only if the second testing had at least one positive replicate with a Cq value <40. If the IAC failed in qPCR, the sample was re-extracted; if the IAC failed again after re-extraction, the sample was excluded from analyses. In all cases of re-testing, the Cq value of the re-test was used in the analysis. We tested for inhibition using the *N. americanus* assay, described in detail in Supplemental Information (Inhibition Testing).

#### Droplet digital PCR

A subset of 50 randomly selected soil DNA aliquots from Benin, India and Kenya were tested for *N. americanus, A. lumbricoides,* and *T. trichiura* by droplet digital PCR (ddPCR) at Christian Medical College, Vellore. All primers and probes were identical to those described above for qPCR and reactions were performed using the QX200 Droplet Digital^TM^ PCR system (Bio-Rad). The ddPCR reaction mix for each target assay consisted of 11 µl of 2x ddPCR^TM^ Supermix for Probes (Bio-Rad); primers (250 nM concentrations of each primers for *N. americanus* and 62.5 nM concentrations for *A. lumbricoides* and *T. trichiura*); probes (125 nM concentrations for all assays) and 4 µL of sample DNA, resulting in a final reaction volume of 22 µL. Droplets were generated in the QX200^TM^ droplet generator (Bio-Rad) with 20 µL of the reaction mix and 70 µL of droplet generating oil in an 8 channel DG8 cartridge. Droplets in oil suspensions were transferred to a 96-well semi-skirted ddPCR plate (Bio-Rad) and placed into a C1000 Touch Thermal Cycler (Bio-Rad). Cycling conditions included an initial denaturation step at 95°C for 10 min, followed by 40 cycles of 94°C for 30 sec and 59°C for 1 min. Cycling was followed by a final hold at 98°C for 10 min. Droplets were read automatically by the QX200^TM^ droplet reader (Bio-Rad) and the data was analyzed with the QuantaSoft™ Version 1.7.4 (Bio-Rad). Any sample with 3 or more droplets at least in one well was considered positive.

### 2.6 Statistical Analyses

To evaluate the agreement between ddPCR and qPCR, and between microscopy and qPCR for the detection of STH in soil, we calculated the percent agreement as the number of samples for which the two methods agreed (either positive/positive or negative/negative), divided by the total number of samples tested. We then estimated the Kappa statistic, with asymptotic standard errors and *P-*values using an alpha of 0.05 for statistical significance, to determine whether agreement was poor (κ < 0), slight (0.01-0.20), fair (0.21-0.40), moderate (0.41-0.60), substantial (0.61-0.80), or perfect (0.81-1.00).^28^

We estimated bivariate associations between characteristics of soil samples and detection of STH in soil samples using logistic regression models. Outcome variables were presence/absence for each STH target in each soil sample, detected via qPCR. Soil characteristics of interest included soil sampling location, soil type, moisture content, shade/sun, presence of feces, and pH; these variables are further described in Supporting Information (Table S3). We used generalized estimating equations (GEE) with an exchangeable working correlation to estimate robust standard errors and adjust for repeated soil samples at the household level.

We also estimated associations between soil STH prevalence and stool STH prevalence at matched households for each STH target, detected by qPCR. Soil characteristic variables screened for associations with STH in soil were included as covariates in these models if associations were statistically significant, using a cutoff of *p* < 0.20. Outcome variables were presence/absence for each STH target at the household level (i.e., whether any stool sample from that household was positive). For this analysis, we included only households where both soil and stool samples were successfully collected. We also only included STH targets with a household-level stool prevalence of >5% at a given study site to avoid positivity assumption violations due to low outcome prevalence. We report both unadjusted and adjusted associations, where adjusted models included covariates after variable selection for each STH outcome, described above. Study site (country) was also included as a covariate in all adjusted models. We used GEE with an exchangeable working correlation and a Poisson distribution due to the zero-inflated natured of the outcome data to avoid model convergence issues. We report measures of association as odds ratios (OR) with 95% confidence intervals (CI). We used R (V 1.0.143) for all tables, figures, and statistical analyses using packages *tidyr, arsenal, ggplot2, vcd,* and *geepack.^29–34^*

## 3. Results & Discussion

### 3.1 Household and soil sample characteristics

In total, we analyzed 478 soil samples and 669 stool samples for STH across 322 households in Benin, India, and Kenya. Household drinking water sources varied by country, though most households had access to an improved drinking water source (Table 1). Public taps/standpipes were one of the most common water sources in Benin (61%), India (39%), and Kenya (69%). Other common water sources included unprotected dug wells in rural Benin (24%) and tube wells or boreholes in urban Kenya (21%); nearly half of households in India had piped water into the household (49%), compared to almost no households in Benin or Kenya (Table 1).

**Table 1.** Household characteristics in each study site and overall.

|  | Benin<br>(rural)<br>n (%) | India<br>(rural)<br>n (%) | Kenya<br>(urban)<br>n (%) | Total<br>n (%) |
| --- | --- | --- | --- | --- |
| Total Households | 104 (100) | 99 (100) | 119 (100) | 322 (100) |
| Total Soil Samples |  |  |  |  |
| *Household entrance* | 104 (100.0) | 99 (100) | 117 (98.3) | 320 (99.4) |
| *Household water source* | 56 (53.8) | 54 (54.5) | 48 (40.2) | 158 (49.0) |
| Drinking water collection location |  |  |  |  |
| *Directly from a filter* | 0 (0.0) | 0 (0.0) | 5 (4.2) | 5 (1.6) |
| *Directly from storage container* | 52 (50.0) | 91 (91.9) | 114 (95.8) | 257 (79.8) |
| *Directly from water source* | 52 (50.0) | 8 (8.1) | 0 (0.0) | 60 (18.6) |
| Improved drinking water source |  |  |  |  |
| *Public tap/standpipe* | 60 (61.2) | 35 (38.9) | 81 (69.2) | 176 (57.7) |
| *Tube well or borehole* | 7 (7.1) | 1 (1.1) | 24 (20.5) | 32 (10.5) |
| *Piped into dwelling* | 1 (1.0) | 44 (48.9) | 0 (0.0) | 45 (14.8) |
| *Piped to yard/plot* | 5 (5.1) | 6 (6.7) | 7 (6.0) | 18 (5.9) |
| *Protected dug well* | 0 (0.0) | 4 (4.4) | 0 (0.0) | 4 (1.3) |
| Unimproved drinking water source |  |  |  |  |
| *Unprotected dug well* | 23 (23.5) | 0 (0.0) | 0 (0.0) | 23 (7.5) |
| *Cart with small tank or tanker truck* | 0 (0.0) | 0 (0.0) | 4 (3.4) | 4 (1.3) |
| *Unprotected spring* | 2 (2.0) | 0 (0.0) | 0 (0.0) | 2 (0.7) |
| *Other* | 0 (0.0) | 0 (0.0) | 1 (0.9) | 1 (0.3) |
| *N-Miss* | 6 | 9 | 2 | 17 |
| Household drinking water treatment |  |  |  |  |
| *No* | 96 (95.0) | 79 (86.8) | 97 (82.9) | 272 (88.0) |
| *Yes* | 5 (5.0) | 12 (13.2) | 20 (17.1) | 37 (12.0) |
| *N-Miss* | 3 | 8 | 2 | 13 |
| Water storage container |  |  |  |  |
| *Drum (metal/plastic) with lid* | 13 (12.9) | 5 (5.5) | 0 (0.0) | 18 (5.9) |
| *Drum (metal/plastic) without lid* | 2 (2.0) | 0 (0.0) | 0 (0.0) | 2 (0.7) |
| *Jerrycan (metal/plastic)* | 6 (5.9) | 0 (0.0) | 65 (56.5) | 71 (23.1) |
| *Plastic tub or bucket with lid* | 9 (8.9) | 0 (0.0) | 34 (29.6) | 43 (14.0) |
| *Plastic tub or bucket without lid* | 0 (0.0) | 0 (0.0) | 1 (0.9) | 1 (0.3) |
| *Water or cooking pot (plastic/metal/clay)* | 71 (70.3) | 86 (94.5) | 8 (7.0) | 165 (53.7) |
| *Water storage vessel with lid* | 0 (0.0) | 0 (0.0) | 4 (3.5) | 4 (1.3) |
| *Other* | 0 (0.0) | 0 (0.0) | 3 (2.6) | 3 (1.0) |
| *N-Miss* | 3 | 8 | 4 | 15 |
| Water use method |  |  |  |  |
| *Container/glass dipped into water container* | 99 (98.0) | 91 (100.0) | 29 (24.8) | 219 (70.9) |
| *Ladle used to obtain water* | 0 (0.0) | 0 (0.0) | 7 (6.0) | 7 (2.3) |
| *Water poured from container* | 2 (2.0) | 0 (0.0) | 75 (64.1) | 77 (24.9) |
| *Water poured from tap/handpump* | 0 (0.0) | 0 (0.0) | 6 (5.1) | 6 (1.9) |
| *N-Miss* | 3 | 8 | 2 | 13 |
| Owens poultry | 80 (76.9) | 17 (17.2) | 58 (48.7) | 155 (48.1) |
| Owens dogs | 32 (30.8) | 4 (4.0) | 6 (5.0) | 42 (13.0) |
| Owens ruminants | 58 (56.3) | 42 (42.4) | 2 (1.7) | 102 (31.8) |
| *N-Miss* | 1 | 0 | 0 | 1 |

Of 478 soil samples, 67% were collected from the household entrance while 33% were collected at the household water source (Table 2). Soil samples were most frequently classified as sand (32%) or loamy sand (18%) in Benin, sandy loam (47%) or loam (33%) in India, and loam (17%), sandy loam (14%), clay loam (14%), or sand (14%) in Kenya. Feces was visible near 27% of all soil sampling locations across the different study sites. Soil moisture content was highest on average in Kenya (mean: 22%, standard deviation (SD): 12%), while soil pH was highest in Benin (mean: 8.06, SD: 0.34) (Table 2). Moisture content was also slightly higher in soil collected from household water sources (mean: 13.30, SD: 10.87) compared to soil from the household entrance (mean: 10.22, SD: 11.66) (SI, Table S5.)

**Table 2.** Characteristics of soil samples collected in each study site and overall.

|  | Benin<br>n (%) | India<br>n (%) | Kenya<br>n (%) | Total<br>n (%) |
| --- | --- | --- | --- | --- |
| Total Samples | 160 (100) | 153 (100) | 165 (100) | 478 (100) |
| Sample Type |  |  |  |  |
| *Household entrance soil* | 104 (65.0) | 99 (64.7) | 117 (70.9) | 320 (66.9) |
| *Water source soil* | 56 (35.0) | 54 (35.3) | 48 (29.1) | 158 (33.1) |
| Soil Type A (most stable) | 63 (39.4) | 69 (45.1) | 101 (61.2) | 233 (48.7) |
| *Clay* | 3 (1.9) | 1 (0.7) | 6 (3.7) | 10 (2.1) |
| *Clay loam* | 19 (11.9) | 9 (5.9) | 22 (13.6) | 50 (10.5) |
| *Loam* | 14 (8.8) | 51 (33.3) | 27 (16.7) | 92 (19.4) |
| *Sandy clay* | 2 (1.2) | 0 (0.0) | 0 (0.0) | 2 (0.4) |
| *Sandy clay loam* | 11 (6.9) | 3 (2.0) | 13 (8.0) | 27 (5.7) |
| *Silty clay* | 0 (0.0) | 1 (0.7) | 11 (6.8) | 12 (2.5) |
| *Silty clay loam* | 14 (8.8) | 4 (2.6) | 22 (13.6) | 40 (8.4) |
| Soil Type B | 17 (10.6) | 84 (54.9) | 36 (21.8) | 137 (28.7) |
| *Sandy loam* | 10 (6.2) | 72 (47.1) | 23 (14.2) | 105 (22.0) |
| *Silt loam* | 7 (4.4) | 12 (7.8) | 13 (8.0) | 32 (6.7) |
| Soil Type C (least stable) | 80 (50.0) | 0 (0.0) | 25 (15.2) | 105 (22.0) |
| *Loamy sand* | 29 (18.1) | 0 (0.0) | 3 (1.9) | 32 (6.7) |
| *Sand* | 51 (31.9) | 0 (0.0) | 22 (13.6) | 73 (15.4) |
| Feces visible at sampling location | 44 (27.5) | 32 (20.9) | 39 (23.6) | 115 (24.1) |
| Soil visibly wet | 86 (53.8) | 16 (10.5) | 63 (38.2) | 165 (34.5) |
| Soil in sun |  |  |  |  |
| *Partly sunny* | 23 (14.4) | 1 (0.7) | 91 (55.2) | 115 (24.1) |
| *Shaded* | 28 (17.5) | 3 (2.0) | 18 (10.9) | 49 (10.3) |
| *Sunny* | 109 (68.1) | 149 (97.4) | 56 (33.9) | 314 (65.7) |
| Soil moisture (%) |  |  |  |  |
| *Mean (SD)* | 6.19 (5.55) | 5.00 (7.35) | 21.92 (11.29) | 11.24 (11.48) |
| *Range* | 0.00 – 19.65 | 0.00 – 43.42 | 0.00 – 67.21 | 0.00 – 67.21 |
| Soil pH |  |  |  |  |
| *Mean (SD)* | 8.06 (0.34) | 7.81 (0.28) | 7.88 (0.46) | 7.92 (0.39) |
| *Range* | 6.95 - 9.35 | 6.63 - 8.54 | 5.66 - 9.34 | 5.66 - 9.35 |

### 3.2 STH detection in soil samples by microscopy, qPCR, and ddPCR

When determining LODs for STH in soil samples by qPCR, detection limits varied for each helminth species. Through a series of spiking experiments, we determined our new method has a detection limit of five *A. lumbricoides* eggs per 20 g of soil (0.25 eggs per gram[epg] of soil), two hookworm eggs per 20 g of soil (0.1 epg soil), and ten *T. trichiura* eggs per 20 g of soil (0.5 epg soil).

All extraction blanks (n=14) and non-template control (NTC) wells (n=166) were negative for all target STH qPCR assays. All qPCR plates had detection of positive controls for each assay. IAC spiking results are reported in SI (Internal Amplification Control Results). After removing samples without IAC amplification for analysis, our final dataset included 160 soil samples from 104 households in Benin, 152 soil samples from 99 households in India, and 137 soil samples from 102 households in Kenya (449 total soil samples from 305 households).

Field testing of soil in India, Benin, and Kenya demonstrated that STH DNA is frequently detected in soil from households and drinking water sources. By qPCR detection, the overall prevalence of *A. lumbricoides* was 31%, *T. trichiura* was 3%, and any hookworm species (*N. americanus, A. duodenale,* or *A. ceylanicum)* was 24%. *Ascaris* was the predominant STH in soil samples from Benin (26%) and Kenya (59%), while hookworm was the predominant STH in India (37%) (SI Table S1). qPCR detected up to three different hookworm species, with *N. americanus* predominant in Benin and *A. duodenale* predominant in India and Kenya (SI Table S1). *A. ceylanicum* was only assessed by qPCR in India; all stool samples were negative and one household water source soil sample was positive. Due to the low prevalence, these results are not included in analyses.

In comparisons of qPCR versus ddPCR STH detection in soil samples, there was good agreement between the two approaches. We found 78% agreement for *N. americanus* detection by qPCR and ddPCR, 87% agreement for *A. duodenale,* 84% for *A. lumbricoides,* and 85% for *T. trichiura* across the study sites (Figure 1, SI Table S2). Kappa statistics of agreement indicated statistically significant fair to substantial agreement between qPCR and ddPCR STH detection overall for each species (Figure 1, SI Table S2). ddPCR had slightly better sensitivity. However, we identified several advantages of qPCR over ddPCR, including reduced variability between replicates (SI Table S6), comparable sensitivity, and lower cost and wider availability of equipment.

**Figure 1.**
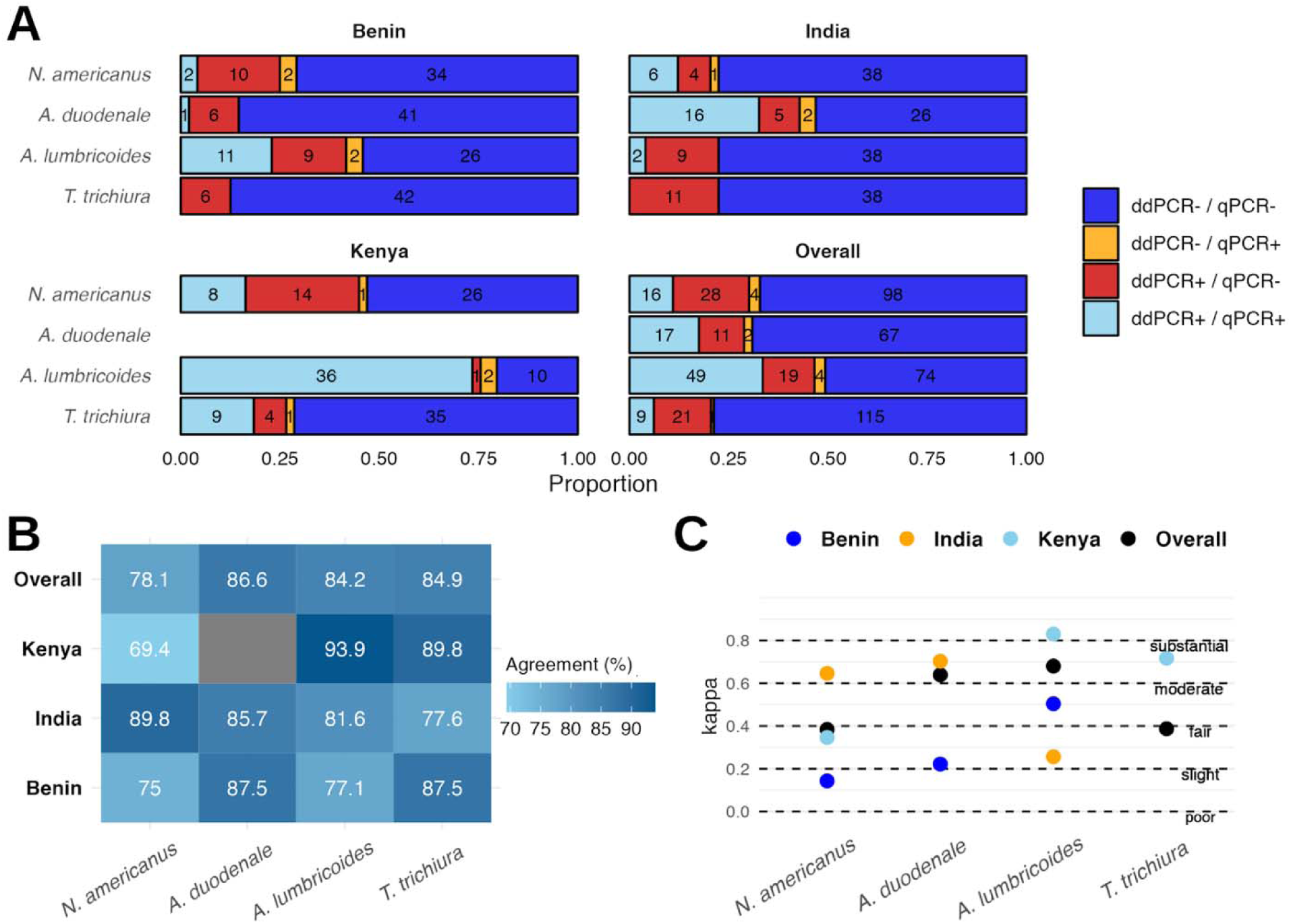
STH detection in soil using qPCR versus ddPCR (A), with percent agreement (B) and Cohen’s Kappa statistic to assess strength of agreement (C). Criteria for positivity by ddPCR was an average of ≥3 positive droplets (2 replicates run).

Agreement between microscopy and qPCR for STH detection in soil was lower than agreement between ddPCR and qPCR. Overall agreement across all study sites was 74% for hookworm, 73% for *Ascaris*, and 81% for *Trichuris* (SI Table S3). Kappa statistics indicated poor agreement, with the highest and only statistically significant Kappa statistic indicating fair agreement for *Ascaris* (SI Table S3). By light microscopy, the prevalence of *Ascaris* was lower than qPCR (20% versus 31%), *Trichuris* was higher (16% versus 3%), and hookworm was substantially lower (6% versus 24%) in soil samples (SI Table S1). In Kenya, *Ascaris* was detected in 30% of soil samples using microscopy but was detected in 63% of samples using qPCR, suggesting higher sensitivity by qPCR (SI Table S3). The prevalence of hookworm in soil was substantially underestimated using microscopy versus qPCR in all three study sites, likely because hookworm degrades during the soil microscopy protocol which takes almost 24 hours to complete. By microscopy, *Trichuris* was detected in 11% and 19% of soil samples in India and Benin, but was not detected in any soil samples in India or Benin via qPCR (SI Table S3). *Trichuris* was also at higher prevalence by microscopy in Kenya compared to qPCR. These results suggest false positive classifications for *Trichuris* by microscopy, which may be other animal-infecting *Trichuris* species rather than human-infecting *T. trichiura.* The discrepancies between prevalence estimates by microscopy and qPCR highlights the challenges associated with identification of human-specific STH species in soil using microscopy and the potential for subjectivity in STH determination. Previous studies of human stool samples have identified better correlation between qPCR and Kato-Katz microscopy results for both *Ascaris* and hookworm.^13^ Our results emphasize the importance in employing molecular methods for soil surveillance of STH to ensure human-specific STH are being monitored.

Soil characteristics associated with detection of any STH species (by qPCR) below a significance threshold of *p*<0.2 included soil type, soil moisture content, sun exposure in the sampling area, and whether there were visible feces near the sampling site. Bivariate associations varied based on the target STH species (Table 3). Samples classified as soil Type C – the least stable soil type including sand and loamy sand – had significantly lower odds of *A. duodenale* detection (OR: 0.18, 95% CI: 0.07, 0.47) (Table 3). In microscopy-based studies, recovery rates of hookworm and other STH ova have been lower in sandy soils compared to clay soils.^35–39^ Molecular methods may improve detection of certain STH in a variety of soil types. Exposure to full sun was associated with lower odds of *A. lumbricoides* detection in soil (OR: 0.17, 95% CI: 0.10, 0.29), while the odds of *A. lumbricoides* detection was two times higher if the soil was visibly wet at the time of sample collection (OR: 2.29, 95% CI:1.44, 3.63) (Table 3). *T. trichiura* detection in soil was more than four times as likely if the soil was visibly wet (OR: 4.37, 95% CI: 1.31, 14.57) (Table 3). Soil moisture content was positively associated with detection of all STH species, as has been found in a previous study in Kenya using microscopy to detect STH.^40^ Given the role of sunlight in desiccation and soil moisture in the growth and activation of STH species, these two variables are potentially important for understanding the role of soil-reservoirs of STH in a community and considerations for soil sampling strategies.

**Table 3.**
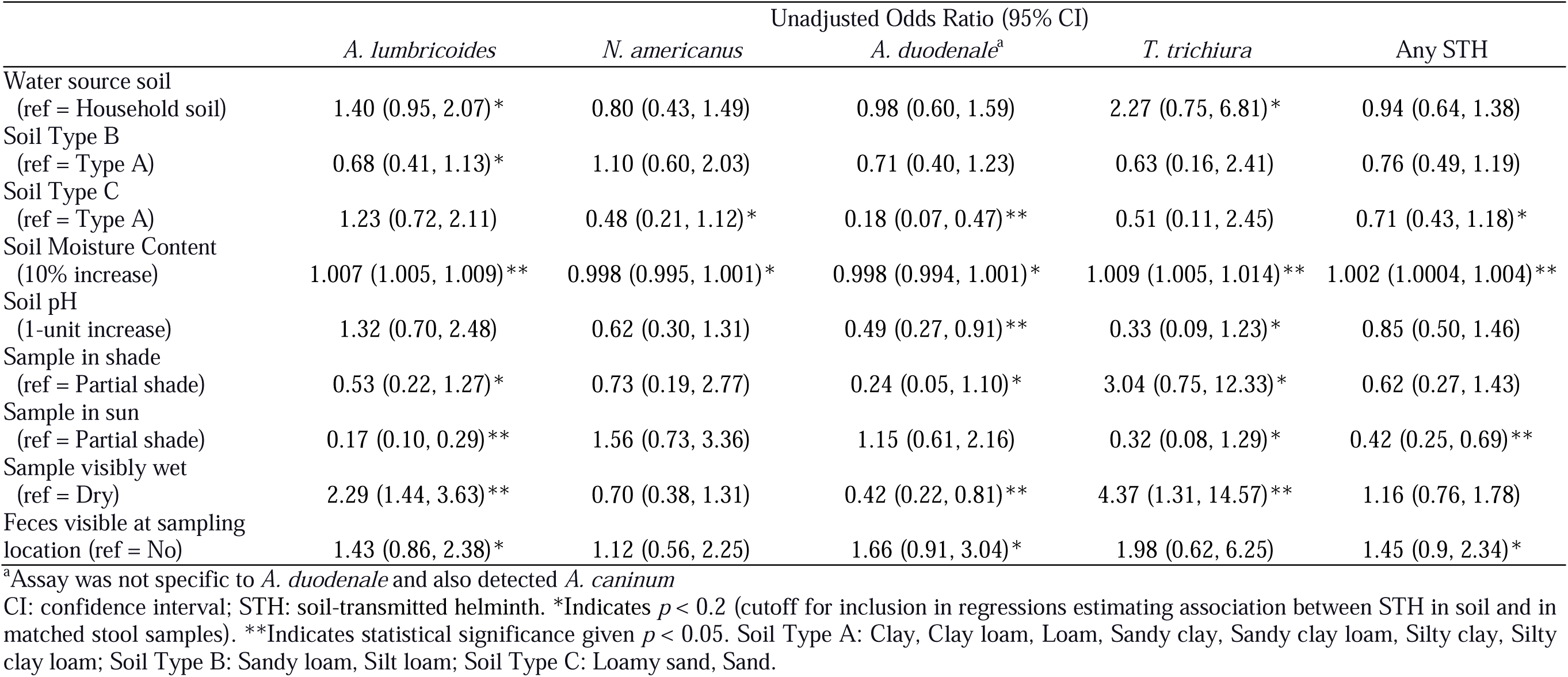
Bivariate associations between soil characteristics and soil-transmitted helminth (STH) detection in soil samples (n=449) by qPCR.

### 3.3 qPCR detection of STH in household-matched soil and stool samples

We assessed STH prevalence in 669 human stool samples in Benin (N=248), India (N=142), and Kenya (N=279). *A. lumbricoides* was detected in individual stool samples from Benin (5%) and Kenya (25%), though it was not detected in any samples in India (SI Table S1). A recent study across 20 counties in western Kenya reported a prevalence of 9.7% (95% CI: 7.5–12.6) for *A. lumbricoides* infections among school children after five rounds of MDA.^41^ Kenya had the highest infection prevalence for *T. trichiura* (5.4%), while India had the highest prevalence of *N. americanus* (19%) (SI Table S1). Though *A. duodenale* was not detected in any stool samples, hookworm prevalence in our study site was higher than previous prevalence estimates in India; a meta-analysis reported a 5% prevalence (95% CI: 0.03, 0.10) for hookworm infection based on data collected from 45,179 participants in India across 46 studies.^42^

Overall, 31.9% of households (n=307) had at least one stool sample that was positive for any given STH species (Table 4). STH were detected more frequently in soil from household water sources (44.5%) and household entrances (46.3%). *A. lumbricoides* was the most frequently detected STH species in soil at household entrances (28.6%), in soil at water sources (34.8%), and in humans (17.6%) (Table 4). STH prevalence was similar in soil from the household entrance and in soil from household water sources across all STH species (Table 4). Many households shared water sources, resulting in fewer soil samples being collected at water sources compared with individual households. Where possible, sampling soil at water sources may be a more efficient strategy for estimating community-level infection prevalence of STH.

**Table 4.** Overall household-level prevalence of soil-transmitted helminths (STH) detected by qPCR in soil and stool across all study sites in Benin, India, and Kenya.

|  | Household Water Source<br>Soil (N=155)<br>n (%) | Household Entrance<br>Soil (N=294)<br>n (%) | Household Stool<sup>a</sup><br>(N=307)<br>n (%) |
| --- | --- | --- | --- |
| *A. lumbricoides* | 54 (34.8) | 84 (28.6) | 54 (17.6) |
| *N. americanus* | 17 (11.0) | 39 (13.3) | 43 (14) |
| *A. duodenale*<sup>b</sup> | 27 (17.4) | 52 (17.7) | 0 (0) |
| *T. trichiura* | 7 (4.52) | 7 (2.38) | 15 (4.89) |
| Any STH | 69 (44.5) | 136 (46.3) | 98 (31.9) |

<sup>a</sup> For stool samples, household-level prevalence was determined based on whether any stool samples (of up to 4) collected from a household were positive for a given STH target.
<sup>b</sup> Assay was not specific to *A. duodenale* and also detected *A. caninum*

In matched stool and soil samples, soil STH profiles via qPCR detection typically reflected stool STH infection profiles across the study sites. *A. lumbricoides* was the most frequently detected STH target in soil samples (62.8%) and in stool from matched households (40.4%) in Kenya (Figure 2). In India, *N. americanus* was most frequently detected in stool (27.1% of households), with a similar detection frequency in soil (17.8%) (Figure 2). We observed that even when human infection prevalence is low, STH DNA can still be detected in soil. For example, *N. americanus* infection prevalence among the 109 households sampled in Kenya was 3.7%, while the prevalence in soil from matched households was 10.2% (Figure 2).

**Figure 2.**
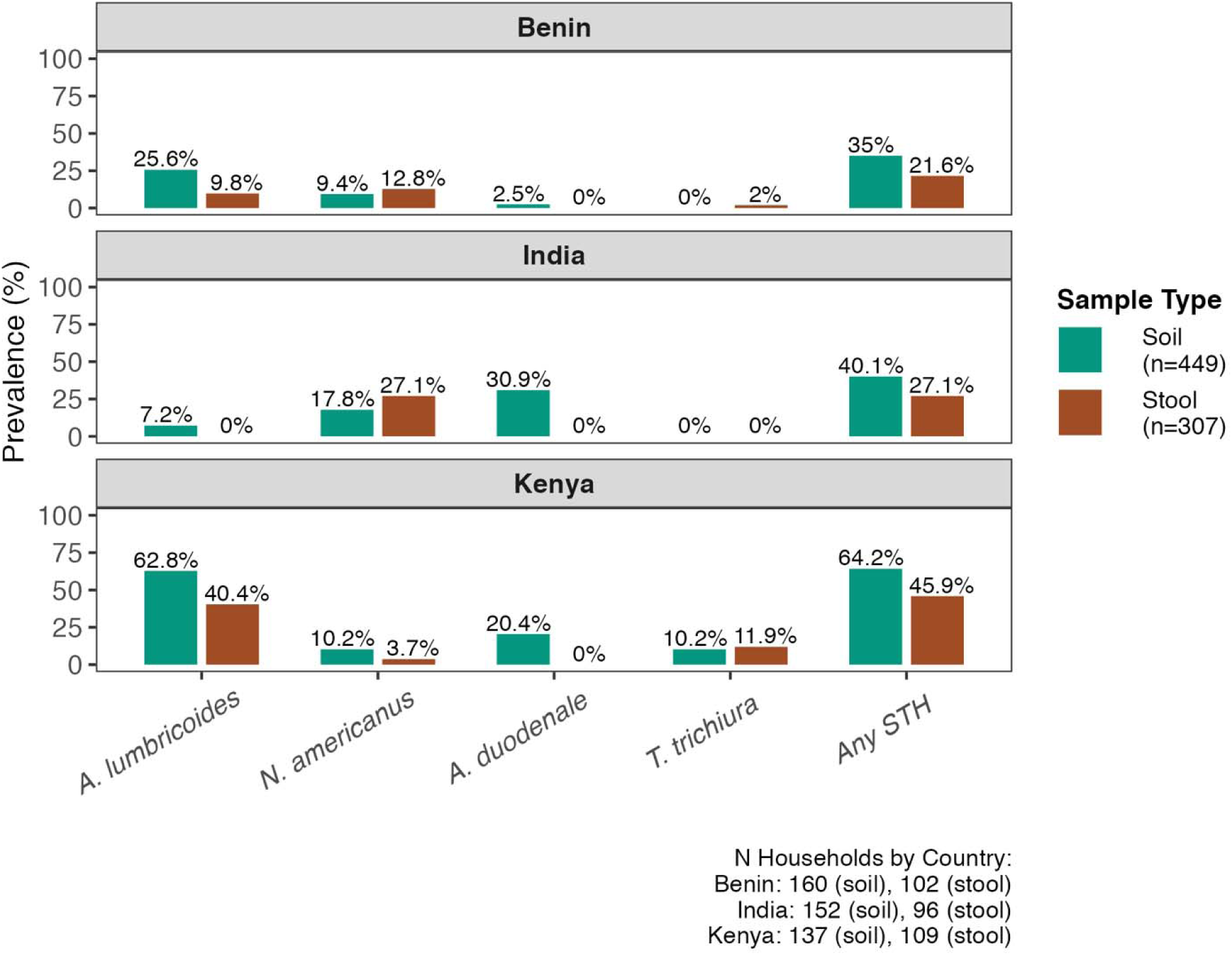
Household-level prevalence of *A. lumbricoides*, *N. americanus*, *A. duodenale, T. trichiura*, and any soil-transmitted helminths (STH) by qPCR detection, stratified by country and sample type. For stool samples, household-level prevalence is determined based on whether any stool samples (of up to 4) collected from a household were positive for a given STH target.

Notably, *A. duodenale* was not detected in any human stool samples but was detected in soil across all three study sites (Figure 2). Sequencing indicated the presence of *A. caninum* – a zoonotic species of *Ancylostoma* which primarily infects dogs – in soil samples testing positive for *Ancylostoma* by qPCR (SI Table S7). This suggested that the qPCR assay used for detection of *A. duodenale* was likely allowing for amplification of *A. caninum*. While the *Ancylostoma* assay initially used in this paper was previously demonstrated to discriminate between *A. duodenale* and *A. ceylanicum*, testing against *A. caninum* did not occur during initial assay development.^14^ This motivated the design of new assays capable of differentiating between *A. duodenale* and *A. caninum* (SI Assay Development Methods).

To ensure that each new assay would not allow for amplification of other *Ancylostoma spp*., other species of STH, human DNA, or common gut flora, optimized assays were tested against 200 pg masses of gDNA from *A. duodenale, A. caninum, A. ceylanicum, T. trichiura, A. lumbricoides, N. americanus, S. stercoralis, S. mansoni, E. coli*, human gDNA, and mixed microbial community gDNA. Both the *A. duodenale* assay and the *A. ceylanicum* assay demonstrated amplification of their intended target but failed to amplify template gDNA from any other species (SI Table S11).

In regression analyses, STH detection in soil was strongly linked to detection of most STH targets in matched household samples (n=290 households after removing samples with failed IAC or missing soil characteristic data) with and without adjustment for soil characteristics. The odds of *A. lumbricoides* detection in stool was 3.74 times higher given detection in matched household soil (aOR: 3.74, 95% CI: 1.99, 7.03) (Figure 3). The odds of *T. trichiura* detection in stool was nearly 10 times higher given detection in matched household soil (aOR: 9.74, 95% CI: 3.31, 28.61), though the estimates were imprecise due to the low prevalence of *T. trichiura* (Figure 3). Hookworm in soil was marginally associated with hookworm in stool from matched households, but only for *N. americanus* (aOR: 1.49, 95% CI: 0.88, 2.52) and the association was not statistically significant (Figure 3). There was no association between *A. duodenale* detection in soil and in stool (aOR: 1.00, 95% CI: 0.75, 1.33) (Figure 3). When considering any STH species, the odds of detection in stool was 1.78 times higher given detection in matched household soil (aOR: 1.78, 95% CI: 1.31, 2.44) (Figure 3). *A. duodenale* was excluded from the regression analysis given the assay’s lack of specificity (based on sequencing results indicating the assay also detected *A. caninum*.

**Figure 3.**
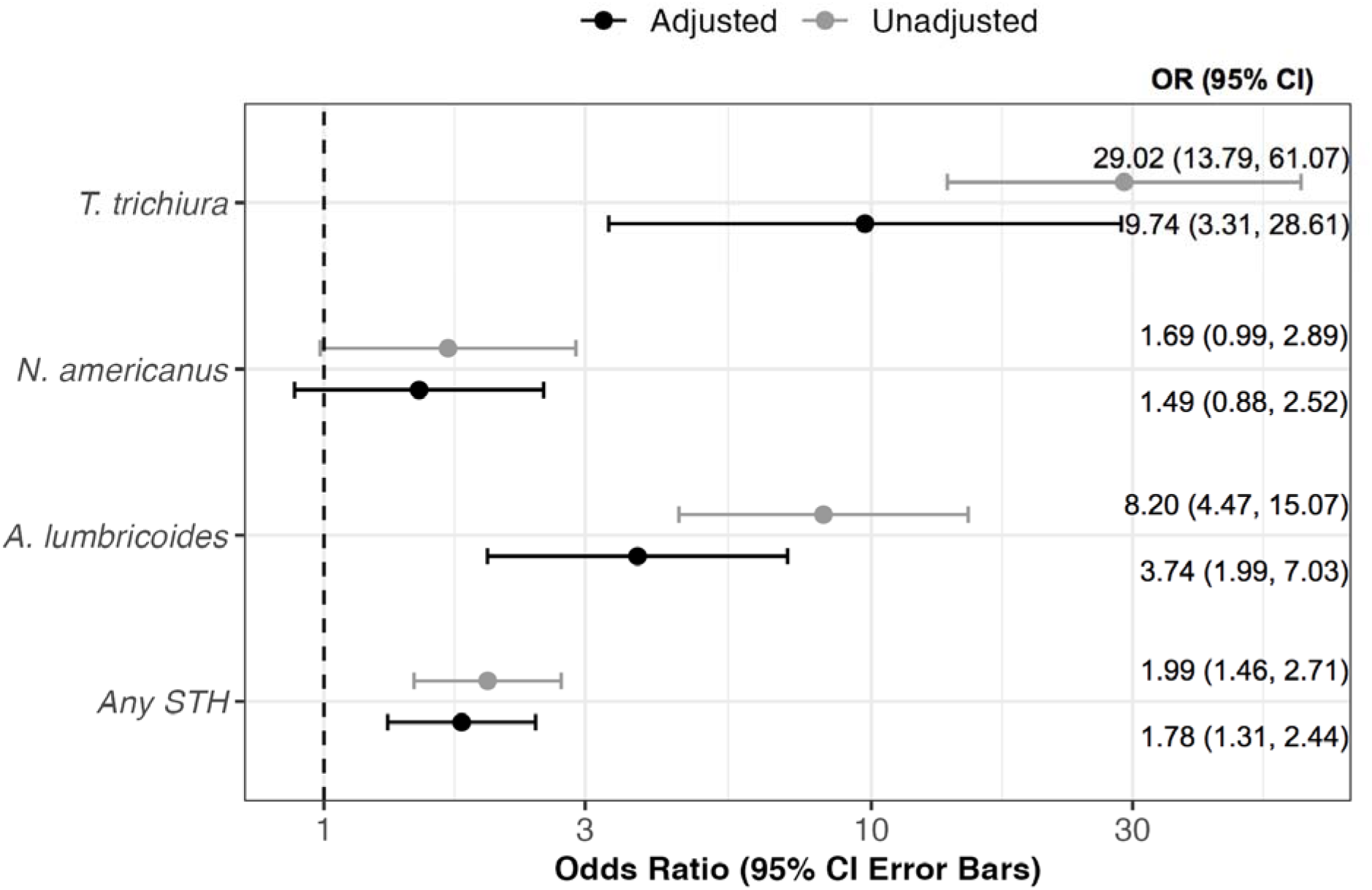
Unadjusted and adjusted associations between soil-transmitted helminth (STH) detection by qPCR in soil and matched human stool samples from 290 households in India, Kenya, and Benin. Points represent odds ratios (OR) with 95% confidence interval (CI) error bars. Adjusted models included covariates associated (p < 0.20) with soil STH detection for each target, and all adjusted models including study site (country). Adjusted model for “Any STH” included variables for sun exposure, soil moisture content, soil type, and whether the sampling area had visible feces nearby; *N. americanus* adjusted model included soil type and moisture content; *A. lumbricoides* adjusted model included sample type, soil type, soil moisture, sun exposure, whether the sampling area was visibly wet, and whether there was visible feces; *T. trichiura* adjusted model included sample type, soil moisture, pH, sun exposure, and whether the sampling area was visibly wet.

To our knowledge, this is the first study to assess the utility of measuring STH in environmental reservoirs by comparing STH prevalence between matched stool and soil samples. The strong univariate (i.e., unadjusted) associations between STH detection in environmental and human samples indicate that environmental sampling could be a useful tool for human STH infection surveillance, even when data on the characteristics of environmental samples are unavailable. Furthermore, considering the comparable results between adjusted and unadjusted models, soil sampling data alone may be sufficient for predicting STH prevalence at the community level, reducing requirements for field surveys or additional observational data.

### 3.4 Implications for Programs and Policy

Through laboratory and field experiments we optimized a method to process and analyze large quantity (up to 20 g) soil samples for STH DNA. Detection of *T. trichiura* was possible at 0.5 epg of soil, *A. lumbricoides* at or above 0.25 epg soil, and *N. americanus* at or above 0.1 epg soil. The method has improved sensitivity over previously published detection limits for light microscopy of soil (1 epg of soil)^18^ and has several strengths. First, direct extraction of DNA from raw soil does not require lengthy flotation or other egg concentration steps that could result in egg loss.^18^ Second, samples can be collected and stored frozen, enabling analysis in batches when convenient. Third, the method is novel in allowing for processing of large quantities of soil (up to 20 g per sample), whereas other soil DNA extraction methods are typically limited to <0.5 g. Fourth, using molecular assays allowed us to ensure we were using assays that are specific to relevant STH species that infect humans. The latter is particularly important for environmental surveillance, as soil samples may contain a variety of animal-specific STH that are not relevant for assessing human infection prevalence. Our results also emphasize the need for ensuring that assays that will be used on environmental samples are comprehensively validated for specificity across all potential animal-infecting helminths that could be present in a study area.

While light microscopy protocols for STH detection in soil can be low-cost and avoid the need for expensive equipment, our results suggest that specificity and sensitivity is limited.^18^ Protocols for concentrating eggs from soil use a series of sieving, flotation, and centrifugation steps, which can be labor intensive, time consuming, and prone to both human error and egg loss. Highly trained and experienced lab technicians are needed to ensure correct identification of STH eggs in soil samples, as samples can contain many types of non-STH nematode eggs. Additionally, hookworm can be too fragile to withstand the processing time required for microscopy (e.g. 24 hours) resulting in false negative samples.^18^ For example, studies in rural Kenya and rural Bangladesh that collected and processed > 2000 soil samples did not detect hookworm in any sample.^43^ In this study, *Trichuris* detected by light microscopy in 11 samples from the India study site were negative by qPCR, suggesting that these samples may have contained a morphologically similar but different species of *Trichuris* (e.g. *Trichuris ovis* that infects goats). Other studies have reported misclassification of human STH through microscopy, with particularly low sensitivity for the detection of hookworm in low prevalence settings.^13,17,44,45^

Environmental surveillance has several benefits over human stool sampling for the estimation of community-level STH prevalence. First, the success rate for sample collecting is significantly higher for soil sampling compared to stool sampling. Stool sampling requires enumerators to visit participating households first to drop off a collection kit, followed by multiple return visits to successfully collect the sample. Soil sampling can be done at a single visit, concurrent with other data collection. At baseline, stool sampling among consenting participants in the DeWorm3 cluster-randomized controlled trial had an 89.8% (6092/6783) success rate in Benin and an 87.3% (6152/7054) success rate in India. In Kenya, the stool sampling success rate for this pilot study was 85%. Notably these stool sampling success rates were achieved through extensive community sensitization and visiting households up to three times for sample retrieval. In contrast, the soil sampling success rate at participating households was 95% (98/103) in India, 100% (106/106) in Benin, and 100% (120/120) in Kenya. The presence of soil at the primary drinking water source was a limiting factor for soil sampling in India; soil was only present at 52% (54/103) of water sources in India, compared to 99% (105/106) in Benin and 93% (111/120) in Kenya. In India, many rural villages in the study site had concrete or cow dung covering much of the area around community water sources. An alternative sampling strategy for concrete surfaces could include sweeping a larger specified area to collect soil. Household soil sampling may be preferred in similar contexts where soil is unavailable at the water source, though sampling at shared household water sources where possible may be a more efficient strategy requiring fewer samples, as STH prevalence in water source soil was comparable to household entrance soil. Moreover, sampling soil at public sites or common spaces like water sources can capture STH circulating in the community rather than at an individual household. Given that MDA programs are delivered at the community level, it is appropriate to also assess STH prevalence through environmental sampling at the community level.

Soil surveillance for STH is a non-intrusive, efficient approach for capturing information on community-level STH infection prevalence. With species-specific Taqman-based qPCR assays, soil sampling can identify the presence of human-infecting STH species in order to target high-burden communities for appropriate interventions. Further validation is needed to optimize the number and location of soil samples needed to predict human infection prevalence within meaningful thresholds (e.g. < 2% prevalence to indicate transmission interruption for a particular species).^19^ Soil surveillance could be a cost-saving tool for monitoring STH prevalence over time, including detecting recrudescence during and after MDA programs. While this study is limited to cross-sectional data, STH prevalence in soil should be compared to STH in humans longitudinally to better differentiate between recrudescence versus persistent environmental contamination due to the long survival times of some STH species.^46^ Data on STH in environmental reservoirs may be used to influence critical programmatic decisions, such as when MDA should be renewed, reduced, or stopped altogether. Longitudinal and localized environmental surveillance of STH transmission – rather than monitoring MDA program coverage – could strengthen longitudinal and localized monitoring efforts for identifying transmission breakpoints and determining whether a sustained break in transmission has been achieved.^47^ Community-level soil surveillance for STH is a feasible, affordable, and efficient strategy for districts and programs looking to enhance their MDA program monitoring as they move beyond morbidity control and towards transmission interruption.

## Data Availability

All data produced in the present work are contained in the manuscript.

## Acknowledgements

We thank the field staff, field supervisors and device managers in India, Benin and Kenya for carrying out data and sample collection and the community members for consenting to participate in this study. This work was funded by the Bill and Melinda Gates Foundation grant numbers OPP1200651 and OPP1129535.

## Notes

### Competing Interest Statement

The authors have declared no competing interest.

### Author Declarations

IRB approval was obtained from Christian Medical College, Vellore (IRB Min no. 10392 dated 08.01.2018), the Ministry of Health in Benin (No. 15/MS/Dc/SGM/DRFMT/CNERS/SA), KEMRI Scientific and Ethics Review Unit (Protocol No. 3823), and Tufts Health Sciences Institutional Review Board (#13205).

### Summary of Updates

Additional details were added on the molecular assays used in the study. Results were also updated based on exclusion of samples in which the internal amplification control did not amplify.

