## Supplemental Information for "Soil surveillance for monitoring soil-transmitted helminth infections: method development and field testing in three countries"

### Assay Development Methods.

In order to develop assays capable of detecting *A. caninum* and *A. duodenale* in a species-specific manner, we utilized a previously described pipeline for next-generation sequencing followed by genomic repeat analysis and characterization (Grant, Pilotte, and Williams 2019). Following sequencing and analysis, putative satellite sequences were chosen as candidate assay targets for both *A. caninum* and *A. duodenale* and both primers and probes were ordered from Integrated DNA Technologies (Coralville, IA). Probes for both assays utilized a 5' 6-FAM fluorophore and were double quenched with ZEN and Iowa Black FQ quenchers (SI, Table S9). Candidate assays then underwent primer optimization to maximize sensitivity of detection. Optimization reactions were performed as previously described and pairings resulting in the lowest Ct values were selected (SI, Table S10) (Pilotte et al. 2016). For *A. duodenale*, 500 nM was determined to be the optimal concentration for both forward and reverse primers. For *A. ceylanicum*, detection was optimal when pairing 125 nM of forward primer with 1000 nM of reverse.

### Inhibition Testing.

We conducted qPCR inhibition testing using a subset of 49 soil samples in India. Samples were run in replicate using both undiluted DNA and diluted (1:2) DNA samples for the *N. americanus* assay. One sample had a positive well when diluted and negative when undiluted, while 5 samples were positive without diluting and negative when diluted. Ct values among the six samples with amplification from both undiluted and diluted DNA did not indicate inhibition.

### Internal Amplification Control Results.

We spiked all samples with an internal amplification control (IAC) plasmid. If the IAC failed to amplify in qPCR, DNA was re-extracted from the raw sample and qPCR for the IAC was rerun. In Benin, the IAC did not fail to amplify in any samples. In India, the IAC failed to amplify in 1 soil sample after re-extraction and rerunning qPCR. In Kenya, we observed higher rates of IAC failure. The IAC failed in 23 samples in initial qPCR runs. Of 40 samples that were re-run (either due to IAC failure or due to single-well amplification of an STH target), 29 samples failed to amplify the IAC. All samples that failed to amplify the IAC after re-extracting DNA and re-running qPCR were removed from the analysis.

### Sanger sequencing of *Ancyclostoma duodenale* qPCR positive soil DNA samples.

An *Ancylostoma* genus-specific nested PCR targeting the ITS 1, 2 and 5.8S region was used in a final volume of 50uL containing 10xPCR buffer, 50mM MgCl2, 10mM dNTP, 10 picomole of each primer, 0.5uL of Taq polymerase, DEPC water and 5uL of DNA.(George et al. 2015) *A. duodenale* positive DNA (obtained from the Wellcome Trust Research Laboratory, Christian Medical College, Vellore) was used as PCR positive control. The PCR was carried out with an initial denaturation at 95°C for 2 min followed by

34 cycles of 95°C for 30 sec (denaturation), 55°C for 30 sec (annealing), 72°C for 30 sec (extension), followed by final denaturation at 72°C for 10 min Agarose gel electrophoresis was carried out using a 1.5% gel in TBE buffer with BioRad PowerPac TM and visualized with BioRad Gel Doc XR system. The PCR product was then purified pre-sequencing with 2uL ExoSap (Cat no.78201.1ML) and 6uL PCR amplicon with heating at 37oC for 5 min followed by 80 o C for 15 min. For the sequencing PCR, the Big dye Terminator Sequencing kit (Cat no.4337455) was used where the forward and reverse sequences were amplified separately in a reaction mixture containing 1uL RR mix, 1.5uL sequence buffer, 3.9uL DEPC water, 1.6uL second round forward/reverse primer and 4uL pre-clean up PCR product. This PCR was carried out at 96o C for 15 sec followed by 25 cycles of 50oC for 20sec, 60oC for 4min, 15oC for 10sec. Post sequencing PCR purification was carried out using Montage sequencing reaction clean up kit (Cat no. LSK09604) followed by sequencing using ABI 3130 Genetic analyzer. Forward and reverse sequence was obtained as AB1 file, contig was prepared with Sequencher 5.4.6 (listed below). All three samples showed greater than 98% identity to *Ancylostoma caninum* (NCBI BLAST, 05 May2021, Table S7).

**Table S1.** Sample-level soil-transmitted helminth (STH) prevalence in soil collected from household entrances and household drinking water sources by microscopy and qPCR, and STH prevalence in stool samples (N=669) by qPCR.

| | | Benin n (%) | India n (%) | Kenya n (%) | Total n (%) |
|---|---|---|---|---|---|
| Soil Samples | **Microscopy (Total Samples)** | 160 | 153 | 165 | 478 |
| | *Ascaris* | 50 (31.2) | 0 (0.0) | 47 (28.5) | 97 (20.3) |
| | *Trichuris* | 31 (19.4) | 17 (11.1) | 30 (18.2) | 78 (16.3) |
| | Hookworm | 1 (0.6) | 24 (15.7) | 2 (1.2) | 27 (5.6) |
| | **qPCR (Total Samples)** | 160 | 152 | 137 | 449 |
| | *Ascaris lumbricoides* | 41 (25.6) | 11 (7.2) | 86 (62.8) | 138 (30.7) |
| | *Trichuris trichiura* | 0 (0.0) | 0 (0.0) | 14 (10.2) | 14 (3.1) |
| | Hookworm | 19 (11.9) | 56 (36.8) | 32 (23.4) | 107 (23.8) |
| | *Necator americanus* | 15 (9.4) | 27 (17.8) | 14 (10.2) | 56 (12.4) |
| | *Ancylostoma duodenale** | 4 (2.5) | 47 (30.9) | 28 (20.4) | 79 (17.6) |
| | *Ancylostoma ceylanicum* | 0 (0.0) | 1 (0.7) | - | 1 (0.3) |
| Stool Samples | **qPCR (Total Samples)** | 248 | 142 | 279 | 669 |
| | *Ascaris lumbricoides* | 13 (5.2) | 0 (0.0) | 69 (24.7) | 82 (12.3) |
| | *Trichuris trichiura* | 2 (0.8) | 0 (0.0) | 15 (5.4) | 17 (2.5) |
| | Hookworm | 15 (6.0) | 27 (19.0) | 6 (2.2) | 48 (7.2) |
| | *Necator americanus* | 15 (6.0) | 27 (19.0) | 6 (2.2) | 48 (7.2) |
| | *Ancylostoma duodenale** | 0 (0.0) | 0 (0.0) | 0 (0.0) | 0 (0.0) |
| | *Ancylostoma ceylanicum* | - | - | - | - |

**A. duodenale* primers also detected *A. caninum* prior to new assay development.

**Table S2.** Agreement of ddPCR and qPCR for the detection of STH in soil among a subset of soil samples from India and Benin (DeWorm3 study sites) randomly selected for ddPCR comparison (n=49).

| | qPCR | ddPCR | | Agreement (%) | Kappa[a] (ASE) | *P*-value |
|---|---|---|---|---|---|---|
| *N. americanus* | | undetected | detected | | | |
| Overall | undetected | 98 | 28 | 78.1 | 0.384 (0.082) | <0.0001 |
| | detected | 4 | 16 | | *fair* | |
| Benin | undetected | 34 | 10 | 75.0 | 0.143 (0.143) | 0.319 |
| | detected | 2 | 2 | | *slight* | |
| India | undetected | 38 | 4 | 89.8 | 0.646 (0.143) | <0.0001 |
| | detected | 1 | 6 | | *substantial* | |
| Kenya | undetected | 26 | 14 | 69.4 | 0.346 (0.115) | 0.003 |
| | detected | 1 | 8 | | *fair* | |
| *A. duodenale* | | undetected | detected | | | |
| Overall | undetected | 67 | 11 | 86.6 | 0.639 (0.089) | <0.0001 |
| | detected | 2 | 17 | | *substantial* | |
| Benin | undetected | 41 | 6 | 87.5 | 0.222 (0.187) | 0.235 |
| | detected | 0 | 1 | | *fair* | |
| India | undetected | 26 | 5 | 85.7 | 0.703 (0.103) | <0.0001 |
| | detected | 2 | 16 | | *substantial* | |
| Kenya | undetected | - | - | - | - | - |
| | detected | - | - | | | |
| *A. lumbricoides* | | undetected | detected | | | |
| Overall | undetected | 74 | 19 | 84.2 | 0.680 (0.06) | <0.0001 |
| | detected | 4 | 49 | | *substantial* | |
| Benin | undetected | 26 | 9 | 77.1 | 0.504 (0.124) | <0.0001 |
| | detected | 2 | 11 | | *moderate* | |
| India | undetected | 38 | 9 | 81.6 | 0.256 (0.15) | 0.087 |
| | detected | 0 | 2 | | *fair* | |
| Kenya | undetected | 10 | 1 | 93.9 | 0.830 (0.0948) | <0.0001 |
| | detected | 2 | 36 | | *perfect* | |
| *T. trichiura* | | undetected | detected | | | |
| Overall | undetected | 115 | 21 | 84.9 | 0.387 (0.097) | <0.0001 |
| | detected | 1 | 9 | | *fair* | |
| Benin | undetected | 42 | 6 | 87.5 | - | - |
| | detected | 0 | 0 | | | |
| India | undetected | 38 | 11 | 77.6 | - | - |
| | detected | 0 | 0 | | | |
| Kenya | undetected | 35 | 4 | 89.8 | 0.717 (0.117) | <0.0001 |
| | detected | 1 | 9 | | *substantial* | |

[a] Cohen's kappa statistic of agreement. <0-poor, 0.01-0.20-slight, 0.21-0.40-fair, 0.41-0.60-moderate, 0.61-0.80-substantial, 0.81-1.00-perfect. ASE: asymptotic standard error.

**Table S3.** Agreement of microscopy and qPCR for the detection of STH in soil, where Kappa statistics indicate agreement from poor, slight, fair, moderate, substantial, to perfect.

| | qPCR Result | Microscopy Results: undetected | Microscopy Results: detected | Agreement[b] (%) | Kappa[c] (ASE) | *P*-value |
|---|---|---|---|---|---|---|
| **Hookworm** | | | | | | |
| Overall | undetected | 323 | 19 | 73.72 | 0.026 (0.038) | 0.4963 |
| | detected | 99 | 8 | | *slight* | |
| Benin | undetected | 141 | 0 | 88.75 | 0.089 (0.083) | 0.2854 |
| | detected | 18 | 1 | | *slight* | |
| India | undetected | 78 | 18 | 55.26 | -0.091 (0.065) | 0.1594 |
| | detected | 50 | 6 | | *poor* | |
| Kenya[a] | undetected | 104 | 1 | 76.64 | 0.032 (0.047) | 0.4968 |
| | detected | 31 | 1 | | *slight* | |
| ***Ascaris*** | | | | | | |
| Overall | undetected | 273 | 38 | 72.61 | 0.289 (0.049) | <0.0001 |
| | detected | 85 | 53 | | *fair* | |
| Benin | undetected | 85 | 34 | 63.13 | 0.098 (0.081) | 0.2279 |
| | detected | 25 | 16 | | *slight* | |
| India | undetected | 141 | 0 | 92.76 | - | - |
| | detected | 11 | 0 | | | |
| Kenya | undetected | 47 | 4 | 61.31 | 0.298 (0.062) | <0.0001 |
| | detected | 49 | 37 | | *fair* | |
| ***Trichuris*** | | | | | | |
| Overall | undetected | 363 | 72 | 81.29 | -0.007 (0.031) | 0.8121 |
| | detected | 12 | 2 | | *poor* | |
| Benin | undetected | 129 | 31 | 80.63 | - | - |
| | detected | 0 | 0 | | | |
| India | undetected | 136 | 16 | 89.47 | - | - |
| | detected | 0 | 0 | | | |
| Kenya | undetected | 98 | 25 | 72.99 | -0.043 (0.071) | 0.5479 |
| | detected | 12 | 2 | | *poor* | |

[a] qPCR assays for hookworm conducted in Kenya included *N. americanus* and *A. duodenale*, whereas qPCR for hookworm detection in Benin and India included assays for *N. americanus*, *A. duodenale,* and *A. ceylanicum.*
[b] Percent agreement is calculated as the number of samples for which qPCR and microscopy results agreed (either positive/positive or negative/negative), divided by the total number of samples tested.
[a] Cohen's kappa statistic of agreement. <0-poor, 0.01-0.20-slight, 0.21-0.40-fair, 0.41-0.60-moderate, 0.61-0.80-substantial, 0.81-1.00-perfect. ASE: asymptotic standard error.

**Table S4.** Soil characteristic variables included in bivariate regressions, where outcome variables were qPCR detection (presence/absence) of soil-transmitted helminths (STH) in soil samples.

| Independent Variables | Treatment Level(s) | Treatment Level Definition | Reference Level | Reference Level Definition |
|---|---|---|---|---|
| Soil Type | Type B | silt loam or sandy loam | Type A | clay, sandy clay, silty clay, loam, sandy clay loam, silty clay loam, clay loam |
| | Type C | loamy sand or sand | | |
| Soil Moisture Content | 10-unit (%) increase | (wet weight – dry weight)/dry weight*100 | No change | - |
| Sample in shade/sun | In shade | Sample fully shaded | Partial shade | Sample partially in shade/sun |
| | In sun | Sample in full sun | | |
| Sample moisture | Wet | Sample visibly wet | Dry | Sample visibly dry |
| Feces near sample | Yes | Feces visible near sample | No | No feces visible near sample |
| pH | 1-unit increase | Mean sample pH | No change | - |
| Sample type | Water source soil | Soil collected at location of water source | Household soil | Soil collected at household entrance |

**Table S5.** Soil sample characteristics by sample type (soil collected from household entrance versus household water source).

| | Household Entrance Soil n (%) | Household Water Source Soil n (%) | Total n (%) |
|---|---|---|---|
| Total Samples | 320 | 158 | 478 |
| Soil Type A | 161 (50.3) | 72 (45.6) | 233 (48.7) |
| *Clay* | 8 (2.5) | 2 (1.3) | 10 (2.1) |
| *Clay loam* | 38 (11.9) | 12 (7.6) | 50 (10.5) |
| *Loam* | 66 (20.8) | 25 (15.9) | 91 (19.2) |
| *Sandy clay* | 0 (0.0) | 2 (1.3) | 2 (0.4) |
| *Sandy clay loam* | 11 (3.5) | 16 (10.2) | 27 (5.7) |
| *Silty clay* | 8 (2.5) | 4 (2.5) | 12 (2.5) |
| *Silty clay loam* | 30 (9.4) | 11 (7.0) | 41 (8.6) |
| Soil Type B | 92 (28.8) | 45 (28.5) | 137 (28.7) |
| *Sandy loam* | 67 (21.1) | 38 (24.2) | 105 (22.1) |
| *Silt loam* | 25 (7.9) | 7 (4.5) | 32 (6.7) |
| Soil Type C | 65 (20.3) | 40 (25.3) | 105 (22.0) |
| *Loamy sand* | 20 (6.3) | 12 (7.6) | 32 (6.7) |
| *Sand* | 45 (14.2) | 28 (17.8) | 73 (15.4) |
| N-Miss (Soil Type) | 2 | 1 | 3 |
| Feces visible at sampling location | 72 (22.5) | 43 (27.2) | 115 (24.1) |
| Soil visibly wet | 110 (34.4) | 55 (34.8) | 165 (34.5) |
| Soil in sun | | | |
| *Partly sunny* | 80 (25.0) | 35 (22.2) | 115 (24.1) |
| *Shaded* | 30 (9.4) | 19 (12.0) | 49 (10.3) |
| *Sunny* | 210 (65.6) | 104 (65.8) | 314 (65.7) |
| Soil moisture (%) | | | |
| *Mean (SD)* | 10.22 (11.66) | 13.30 (10.87) | 11.24 (11.48) |
| *Range* | 0.00 - 67.21 | 0.00 - 49.78 | 0.00 - 67.21 |
| Soil pH | | | |
| *Mean (SD)* | 7.92 (0.41) | 7.92 (0.33) | 7.92 (0.39) |
| *Range* | 5.79 - 9.35 | 5.66 - 9.15 | 5.66 - 9.35 |

**Table S6.** Agreement in soil-transmitted helminth detection between duplicate wells for a subset of samples randomly selected for ddPCR and qPCR.

| | Samples with discrepant replicates (%) | |
|---|---|---|
| | **ddPCR (N=50)** | **qPCR (N=50)** |
| *Ascaris lumbricoides* | | |
| India | 9 (18) | 1 (2) |
| Benin | 11 (22) | 3 (6) |
| Kenya | 2 (4) | 3 (6) |
| *Trichuris trichiura* | | |
| India | 9 (18) | 0 (0) |
| Benin | 7 (14) | 0 (0) |
| Kenya | 4 (8) | 3 (6) |
| *Necator americanus* | | |
| India | 7 (14) | 4 (8) |
| Benin | 10 (20) | 3 (6) |
| Kenya | 18 (36) | 2 (4) |
| *Ancylostoma duodenale* | | |
| India | 7 (14) | 3 (6) |
| Benin | 5 (10) | 0 (0) |
| Kenya | - | - |

**Table S7.** Sanger sequencing results from soil samples (n=4) collected in India that were positive for *A. duodenale* based on qPCR. Raw data was analyzed using Sequencher V 5.4.6.

| Soil Sample | *A. duodenale* qPCR Ct | Sanger sequencing using ITS primers (% identity) | Amplicons |
|---|---|---|---|
| 1 | 30.33 | *A. caninum* (99.26%) | TGGGAGTATCGCCCCCCGTTACAGCCCTACGTAGGTGTCTATGTGCACAAGAGTCGTTACTGGGTGGCGG<br>CAATGATTGCTGTGTGAAGTTCGCGTTTCGCTGAGCTTTAGACTTGATGAGCATTGCATGAATGCCGCCTT<br>ACTGCTTGTGTTGGTGGTTGAGCATTAGGCTAACGCCTGATGCGGCACCTGTCTGTCAGGAAACCTTAAT<br>GATCTGCTAACGCGGACGCCAGTACAGCAATAACTTTTTACGTTTAATGTTTGCAGAATCGTGACTTCACG<br>TCACAATCGACTAGCTTCAGCGATGGATCGGTCGATTCGCGTATCGATGAAAAACGCAGCTAGCTGCGTT<br>ATTTACCACGAATTGCAGACGCTTATAGTGGTGAAATTTTGAACGCATA |
| 2 | 31.2 | *A. caninum* (99.25%) | GGGAGTATCGCCCCCCGTTACAGCCCTACGTAAGGTGTCTATGTGCAGCAAGAGTCGTTACTGGGTGGCG<br>GCAATGATTGCTGTGTGAAGTTCGCGTTTCGCTGAGCTTTAGACTTGATGAGCATTGCATGAATGCCGCCT<br>TACTGCTTGTGTTGGTGGTTGAGCATTAGGCTAACGCCTGATGCGGCACCTGTCTGTCAGGAAACCTTAAT<br>GATCTGCTAACGCGGACGCCAGTACAGCAATAACTTTTTACGTTTAATGTTTGCAGAATCGTGACTTCACG<br>TCACAATCGACTAGCTTCAGCGATGGATCGGTCGATTCGCGTATCGATGAAAAACGCAGCTAGCTGCGTT<br>ATTTACCACGAATTGCATACGCTTAGAGTGGTGAAATTTTGAACGCATA |
| 3 | 33.72 | *A. caninum* (99.26%) | GAGTATCKCCCCCCTGTTGGGAGTATCGCCCCCCGTTACAGCCCTATGTAAGGTGTCTATGTGCAGCAAG<br>AGTCGTTACTGGGTGGCGGCAATGATTGCTGTGTGAAGTTCGCGTTTCGCTGAGCTTTAGACTTGATGAGC<br>ATTGCATGAATGCCGCCTTACTGCTTGTGTTGGTGGTTGAGCATTAGGCTAACGCCTGATGCGTCACCTGT<br>CTGTCAGGAAACCTTAATGATCTGCTAACGCGGACGCCAGTACAGCAATAACTTTTTACGTTTAATGTTTG<br>CAGAATCGTGACTTCACGTCACAATCGACTAGCTTCAGCGATGGATCGGTCGATTCGCGTATCGATGAAA<br>AACGCAGCTAGCTGCGTTATTTACCACGAATTGCAGACGCTTAGAGTGGTGAAATTTTGAACGCATA |
| 4 | 30.84 | *A. caninum* (98.42%) | GTTGGGAGTATCGCCCCCCGTTACAGCCGTACGTAAGGTGTCTATGTGCAGCAAGAGTCGTTGCTGGGTG<br>GCGGCAGTGATTGCTGTGTGAAGTTCGCGTTTCGCTGAGCTTTAGACTTGATGAGCATTGCATGAATGCCB<br>CCTTACTGCTTGTGTTGGTGGTTGAGCATTAGGCTAACGCCTGATGCGGCACCTGTCTGTCAGGAAACCTT<br>AATGATCTGCTAACGCGGACGCCAGTACAGCAATAACTTTTTACGTTTAATGTTTGCAGAATCGTGACTTC<br>ACGTCACAATCGACTAGCTTCAGCGATGGATCGGTCGATTCGCGTATCGATGAAAAACGCAGCTAGCTGC<br>GTTATTTACCACGAATTGCAGACGCTTT |
| Positive Control | 21.16 | *A. duodenale* (100%) | TGTGGTTCAGGGTTGTTACGTGTGTTTCAGGCCTGTTGGGAGTATCGCCCCCCGTTATAGCCCTACGTAAG<br>GTGTCTATGTGCAGCAAGAGTCGTTACTGGGTGGCGGCAGTGATTGCTGTGCGAAGTTCGCGTTTCGCTG<br>AGCTTTAACTTGATGAGCATTGCATGAATGCCGCCTTACTGCTTGTGTTGGTGGTTGAGCATTAGGCTAAC<br>GCCTGATGCGGCACCTGTCTGTCAGGAAACCTTAATGATCTGCTAACGCGGACGCCAGTACAGCAATAAC<br>TTTTTACGTTTAATGTTTGCAGAATCGTGACTTTATGTCACAATCGACTAGCTTCAGCGATGGATCGGTCG<br>ATTCGCGTATCGATGAAAAACGCAGCTAGCTGCGTTATTTACCC |

**Table S8.** Primer and probe sequences used in three-country field study for detection of soil transmitted helminths via qPCR.

| Oligo | Sequence (5' - 3') |
|---|---|
| *A. duodenale* probe | /56-FAM/TGA CAG TGT/ZEN/GTC ATA CTG TGG AAA/3IABkFQ/ |
| *A. duodenale* fwd | GTA TTT CAC TCA TAT GAT CGA GTG TTC |
| *A. duodenale* rev | GTT TGA ATT TGA GGT ATT TCG ACC A |
| *A. ceylanicum* probe | /56-FAM/CGG TGA AAG/ZEN/CTT TGC GTT ATT GCG A /3IABkFQ/ |
| *A. ceylanicum* fwd | CAA ATA TTA CTG TGC GCA TTT AGC |
| *A. ceylanicum* rev | GCG AAT ATT TAG TGG GTT TAC TGG |
| *N. americanus* probe | /56-FAM/CC CGA TTT G/ZEN/A GCT GAA TTG TCA AA/3IABkFQ/ |
| *N. americanus* fwd | CCA GAA TCG CCA CAA ATT GTA T |
| *N. americanus* rev | GGG TTT GAG GCT TAT CAT AAA GAA |
| *A. lumbricoides* probe | /56-FAM/TC TGT GCA T/ZEN/T ATT GCT GCA ATT GGG A/3IABkFQ/ |
| *A. lumbricoides* fwd | CTT GTA CCA CGA TAA AGG GCA T |
| *A. lumbricoides* rev | TCC CTT CCA ATT GAT CAT CGA ATA A |
| *T. trichiura* probe | /56-FAM/TT TGC GGG C/ZEN/G AGA ACG GAA ATA TT/3IABkFQ/ |
| *T. trichiura* fwd | GGC GTA GAG GAG CGA TTT |
| *T. trichiura* rev | TAC TAC CCA TCA CAC ATT AGC C |
| IAC fwd | CTA ACC TTC GTG ATG AGC AAT CG |
| IAC rev | GAT CAG CTA CGT GAG GTC CTA C |
| IAC probe | /56-FAM/AGC TAG TCG/ZEN/ATG CAC TCC AGT CCT CCT/3IABkFQ/ |

IAC: Internal amplification control

**Table S9.** New primer and probe sequences for species-specific detection of *A. duodenale* versus *A. caninum* via qPCR.

| Oligo | Sequence (5' - 3') |
|---|---|
| *A. duodenale* probe | /56-FAM/TGA CAG TGT/ZEN/GTC ATA CTG TGG AAA/3IABkFQ/ |
| *A. duodenale* fwd | TTT GGA TTT GCA GCA GTA TGA C |
| *A. duodenale* rev | CTT CAA ATT CTA ACA GTT GGG TGT A |
| *A. caninum* probe | /56-FAM/TCG TTC GTA/ZEN/TGA GTG AAA CAC ACA CA/3IABkFQ/ |
| *A. caninum* fwd | AGT ATG ACA CAC TGT CAG AAC AC |
| *A. caninum* rev | TGG TCG AAA TAC CTC AAA TTC AAA C |

**Table S10.** Results from primer optimization of candidate assays for *A. duodenale* and *A. caninum.*

| *A. duodenale* | | | | *A. caninum* | | | |
|---|---|---|---|---|---|---|---|
| **Sample Name** | **Ct** | **Ct Mean** | **Ct SD** | **Sample Name** | **Ct** | **Ct Mean** | **Ct SD** |
| 62.5F/62.5R | 25.718 | 25.990 | 0.443 | 62.5F/62.5R | 20.713 | 20.667 | 0.126 |
| 62.5F/62.5R | 26.501 | 25.990 | 0.443 | 62.5F/62.5R | 20.525 | 20.667 | 0.126 |
| 62.5F/62.5R | 25.752 | 25.990 | 0.443 | 62.5F/62.5R | 20.763 | 20.667 | 0.126 |
| 125F/62.5R | 25.042 | 25.130 | 0.372 | 62.5F/125R | 20.747 | 20.644 | 0.090 |
| 125F/62.5R | 24.810 | 25.130 | 0.372 | 62.5F/125R | 20.606 | 20.644 | 0.090 |
| 125F/62.5R | 25.537 | 25.130 | 0.372 | 62.5F/125R | 20.579 | 20.644 | 0.090 |
| 250F/62.5R | 24.930 | 24.891 | 0.178 | 62.5F/250R | 20.980 | 20.861 | 0.113 |
| 250F/62.5R | 24.697 | 24.891 | 0.178 | 62.5F/250R | 20.754 | 20.861 | 0.113 |
| 250F/62.5R | 25.047 | 24.891 | 0.178 | 62.5F/250R | 20.848 | 20.861 | 0.113 |
| 500F/62.5R | 24.893 | 24.795 | 0.122 | 62.5F/500R | 21.123 | 20.634 | 0.440 |
| 500F/62.5R | 24.658 | 24.795 | 0.122 | 62.5F/500R | 20.270 | 20.634 | 0.440 |
| 500F/62.5R | 24.833 | 24.795 | 0.122 | 62.5F/500R | 20.509 | 20.634 | 0.440 |
| 62.5F/125R | 24.316 | 24.670 | 0.326 | 62.5F/1000R | Negative | | |
| 62.5F/125R | 24.957 | 24.670 | 0.326 | 62.5F/1000R | Negative | | |
| 62.5F/125R | 24.736 | 24.670 | 0.326 | 62.5F/1000R | Negative | | |
| 125F/125R | 24.444 | 24.855 | 0.429 | 125F/62.5R | 21.131 | 21.089 | 0.085 |
| 125F/125R | 24.820 | 24.855 | 0.429 | 125F/62.5R | 21.144 | 21.089 | 0.085 |
| 125F/125R | 25.300 | 24.855 | 0.429 | 125F/62.5R | 20.991 | 21.089 | 0.085 |
| 250F/125R | 24.467 | 24.417 | 0.243 | 125F/125R | 21.165 | 20.822 | 0.308 |
| 250F/125R | 24.153 | 24.417 | 0.243 | 125F/125R | 20.737 | 20.822 | 0.308 |
| 250F/125R | 24.632 | 24.417 | 0.243 | 125F/125R | 20.566 | 20.822 | 0.308 |
| 500F/125R | 24.315 | 24.239 | 0.187 | 125F/250R | 21.021 | 20.671 | 0.332 |
| 500F/125R | 24.377 | 24.239 | 0.187 | 125F/250R | 20.633 | 20.671 | 0.332 |
| 500F/125R | 24.026 | 24.239 | 0.187 | 125F/250R | 20.360 | 20.671 | 0.332 |
| 62.5F/250R | 24.603 | 24.600 | 0.148 | 125F/500R | 20.997 | 20.869 | 0.112 |
| 62.5F/250R | 24.451 | 24.600 | 0.148 | 125F/500R | 20.821 | 20.869 | 0.112 |
| 62.5F/250R | 24.747 | 24.600 | 0.148 | 125F/500R | 20.790 | 20.869 | 0.112 |
| 125F/250R | 24.387 | 24.529 | 0.171 | 125F/1000R | 20.816 | 20.622 | 0.201 |
| 125F/250R | 24.719 | 24.529 | 0.171 | 125F/1000R | 20.636 | 20.622 | 0.201 |
| 125F/250R | 24.480 | 24.529 | 0.171 | 125F/1000R | 20.415 | 20.622 | 0.201 |
| 250F/250R | 24.398 | 24.355 | 0.171 | 250F/62.5R | 21.484 | 21.091 | 0.340 |
| 250F/250R | 24.167 | 24.355 | 0.171 | 250F/62.5R | 20.900 | 21.091 | 0.340 |
| 250F/250R | 24.500 | 24.355 | 0.171 | 250F/62.5R | 20.889 | 21.091 | 0.340 |
| 500F/250R | 24.120 | 24.162 | 0.166 | 250F/125R | 20.875 | 20.645 | 0.246 |
| 500F/250R | 24.345 | 24.162 | 0.166 | 250F/125R | 20.676 | 20.645 | 0.246 |
| 500F/250R | 24.021 | 24.162 | 0.166 | 250F/125R | 20.385 | 20.645 | 0.246 |
| 62.5F/500R | 24.273 | 24.561 | 0.776 | 250F/250R | 20.767 | 20.774 | 0.046 |
| 62.5F/500R | 23.970 | 24.561 | 0.776 | 250F/250R | 20.731 | 20.774 | 0.046 |
| 62.5F/500R | 25.440 | 24.561 | 0.776 | 250F/250R | 20.823 | 20.774 | 0.046 |
| 125F/500R | 23.877 | 24.585 | 0.633 | 250F/500R | 20.998 | 21.069 | 0.459 |
| 125F/500R | 24.786 | 24.585 | 0.633 | 250F/500R | 21.560 | 21.069 | 0.459 |
| 125F/500R | 25.094 | 24.585 | 0.633 | 250F/500R | 20.651 | 21.069 | 0.459 |
| 250F/500R | 24.196 | 24.288 | 0.110 | 250F/1000R | 20.880 | 20.679 | 0.175 |
| 250F/500R | 24.410 | 24.288 | 0.110 | 250F/1000R | 20.574 | 20.679 | 0.175 |
| 250F/500R | 24.257 | 24.288 | 0.110 | 250F/1000R | 20.582 | 20.679 | 0.175 |

***Table S10 cont.***

| *A. duodenale* | | | | *A. caninum* | | | |
|---|---|---|---|---|---|---|---|
| **Sample Name** | **Ct** | **Ct Mean** | **Ct SD** | **Sample Name** | **Ct** | **Ct Mean** | **Ct SD** |
| 500F/500R | 23.827 | 23.939 | 0.112 | 500F/62.5R | 21.587 | 21.309 | 0.241 |
| 500F/500R | 23.939 | 23.939 | 0.112 | 500F/62.5R | 21.192 | 21.309 | 0.241 |
| 500F/500R | 24.052 | 23.939 | 0.112 | 500F/62.5R | 21.148 | 21.309 | 0.241 |
| 62.5F/1000R | 24.242 | 24.608 | 0.319 | 500F/125R | 21.077 | 21.075 | 0.170 |
| 62.5F/1000R | 24.760 | 24.608 | 0.319 | 500F/125R | 21.244 | 21.075 | 0.170 |
| 62.5F/1000R | 24.823 | 24.608 | 0.319 | 500F/125R | 20.904 | 21.075 | 0.170 |
| 125F/1000R | 24.905 | 24.410 | 0.494 | 500F/250R | 20.982 | 20.796 | 0.168 |
| 125F/1000R | 24.408 | 24.410 | 0.494 | 500F/250R | 20.753 | 20.796 | 0.168 |
| 125F/1000R | 23.918 | 24.410 | 0.494 | 500F/250R | 20.655 | 20.796 | 0.168 |
| 250F/1000R | 24.406 | 24.208 | 0.378 | 500F/500R | 21.392 | 21.299 | 0.093 |
| 250F/1000R | 23.772 | 24.208 | 0.378 | 500F/500R | 21.206 | 21.299 | 0.093 |
| 250F/1000R | 24.446 | 24.208 | 0.378 | 500F/500R | 21.301 | 21.299 | 0.093 |
| 500F/1000R | 23.966 | 24.139 | 0.162 | 500F/1000R | 21.011 | 20.788 | 0.193 |
| 500F/1000R | 24.163 | 24.139 | 0.162 | 500F/1000R | 20.664 | 20.788 | 0.193 |
| 500F/1000R | 24.288 | 24.139 | 0.162 | 500F/1000R | 20.691 | 20.788 | 0.193 |
| 1000F/62.5R | 24.266 | 24.402 | 0.134 | 1000F/62.5R | 21.312 | 21.398 | 0.117 |
| 1000F/62.5R | 24.408 | 24.402 | 0.134 | 1000F/62.5R | 21.352 | 21.398 | 0.117 |
| 1000F/62.5R | 24.534 | 24.402 | 0.134 | 1000F/62.5R | 21.531 | 21.398 | 0.117 |
| 1000F/125R | 24.201 | 24.409 | 0.181 | 1000F/125R | 21.066 | 21.076 | 0.164 |
| 1000F/125R | 24.530 | 24.409 | 0.181 | 1000F/125R | 21.244 | 21.076 | 0.164 |
| 1000F/125R | 24.497 | 24.409 | 0.181 | 1000F/125R | 20.917 | 21.076 | 0.164 |
| 1000F/250R | 24.508 | 24.590 | 0.074 | 1000F/250R | 21.233 | 21.195 | 0.048 |
| 1000F/250R | 24.610 | 24.590 | 0.074 | 1000F/250R | 21.210 | 21.195 | 0.048 |
| 1000F/250R | 24.651 | 24.590 | 0.074 | 1000F/250R | 21.141 | 21.195 | 0.048 |
| 1000F/500R | 24.267 | 24.176 | 0.224 | 1000F/500R | 20.665 | 20.708 | 0.067 |
| 1000F/500R | 24.341 | 24.176 | 0.224 | 1000F/500R | 20.785 | 20.708 | 0.067 |
| 1000F/500R | 23.921 | 24.176 | 0.224 | 1000F/500R | 20.675 | 20.708 | 0.067 |
| 1000F/1000R | 24.602 | 24.270 | 0.470 | 1000F/1000R | 21.197 | 21.204 | 0.054 |
| 1000F/1000R | 23.938 | 24.270 | 0.470 | 1000F/1000R | 21.154 | 21.204 | 0.054 |
| NTC | Negative | | | 1000F/1000R | 21.261 | 21.204 | 0.054 |
| NTC | Negative | | | NTC | Negative | | |
| NTC | Negative | | | NTC | Negative | | |
| NTC | Negative | | | NTC | Negative | | |

**Table S11.** Specificity testing of *A. duodenale* and *A. ceylanicum* qPCR assays.

| Sample Name | Target Name | Task | Ct | Ct Mean | Ct SD |
|---|---|---|---|---|---|
| A ceylanicum 200pg | A doud soil | UNKNOWN | Negative | | |
| A ceylanicum 200pg | A doud soil | UNKNOWN | Negative | | |
| A ceylanicum 200pg | A doud soil | UNKNOWN | Negative | | |
| A caninum 200pg | A doud soil | UNKNOWN | Negative | | |
| A caninum 200pg | A doud soil | UNKNOWN | Negative | | |
| A caninum 200pg | A doud soil | UNKNOWN | Negative | | |
| T trichuria 200pg | A doud soil | UNKNOWN | Negative | | |
| T trichuria 200pg | A doud soil | UNKNOWN | Negative | | |
| T trichuria 200pg | A doud soil | UNKNOWN | Negative | | |
| N americanus 200pg | A doud soil | UNKNOWN | Negative | | |
| N americanus 200pg | A doud soil | UNKNOWN | Negative | | |
| N americanus 200pg | A doud soil | UNKNOWN | Negative | | |
| A lumbricoides 200pg | A doud soil | UNKNOWN | Negative | | |
| A lumbricoides 200pg | A doud soil | UNKNOWN | Negative | | |
| A lumbricoides 200pg | A doud soil | UNKNOWN | Negative | | |
| S stercoralis 200pg | A doud soil | UNKNOWN | Negative | | |
| S stercoralis 200pg | A doud soil | UNKNOWN | Negative | | |
| S stercoralis 200pg | A doud soil | UNKNOWN | Negative | | |
| S mansoni 200pg | A doud soil | UNKNOWN | Negative | | |
| S mansoni 200pg | A doud soil | UNKNOWN | Negative | | |
| S mansoni 200pg | A doud soil | UNKNOWN | Negative | | |
| Mixed Microb 200pg | A doud soil | UNKNOWN | Negative | | |
| Mixed Microb 200pg | A doud soil | UNKNOWN | Negative | | |
| Mixed Microb 200pg | A doud soil | UNKNOWN | Negative | | |
| Human 200pg | A doud soil | UNKNOWN | Negative | | |
| Human 200pg | A doud soil | UNKNOWN | Negative | | |
| Human 200pg | A doud soil | UNKNOWN | Negative | | |
| E coli 200pg | A doud soil | UNKNOWN | Negative | | |
| E coli 200pg | A doud soil | UNKNOWN | Negative | | |
| E coli 200pg | A doud soil | UNKNOWN | Negative | | |
| Ad+ 20pg | A doud soil | UNKNOWN | 24.642 | 24.619 | 0.191 |
| Ad+ 20pg | A doud soil | UNKNOWN | 24.417 | 24.619 | 0.191 |
| Ad+ 20pg | A doud soil | UNKNOWN | 24.797 | 24.619 | 0.191 |
| Ad+ 2pg | A doud soil | UNKNOWN | 27.564 | 27.681 | 0.157 |
| Ad+ 2pg | A doud soil | UNKNOWN | 27.859 | 27.681 | 0.157 |
| Ad+ 2pg | A doud soil | UNKNOWN | 27.620 | 27.681 | 0.157 |
| Ad+ 200fg | A doud soil | UNKNOWN | 32.927 | 32.254 | 0.662 |
| Ad+ 200fg | A doud soil | UNKNOWN | 31.603 | 32.254 | 0.662 |
| Ad+ 200fg | A doud soil | UNKNOWN | 32.232 | 32.254 | 0.662 |
| NTC | A doud soil | NTC | Negative | | |
| NTC | A doud soil | NTC | Negative | | |
| NTC | A doud soil | NTC | Negative | | |
| A duodenale 200pg | A caninum | UNKNOWN | Negative | | |
| A duodenale 200pg | A caninum | UNKNOWN | Negative | | |
| A duodenale 200pg | A caninum | UNKNOWN | Negative | | |
| A ceylanicum 200pg | A caninum | UNKNOWN | Negative | | |
| A ceylanicum 200pg | A caninum | UNKNOWN | Negative | | |
| A ceylanicum 200pg | A caninum | UNKNOWN | Negative | | |
| T trichuria 200pg | A caninum | UNKNOWN | Negative | | |

*Table S11 cont.*

| Sample Name | Target Name | Task | Ct | Ct Mean | Ct SD |
|---|---|---|---|---|---|
| T trichuria 200pg | A caninum | UNKNOWN | Negative | | |
| T trichuria 200pg | A caninum | UNKNOWN | Negative | | |
| N americanus 200pg | A caninum | UNKNOWN | Negative | | |
| N americanus 200pg | A caninum | UNKNOWN | Negative | | |
| N americanus 200pg | A caninum | UNKNOWN | Negative | | |
| A lumbricoides 200pg | A caninum | UNKNOWN | Negative | | |
| A lumbricoides 200pg | A caninum | UNKNOWN | Negative | | |
| A lumbricoides 200pg | A caninum | UNKNOWN | Negative | | |
| S stercoralis 200pg | A caninum | UNKNOWN | Negative | | |
| S stercoralis 200pg | A caninum | UNKNOWN | Negative | | |
| S stercoralis 200pg | A caninum | UNKNOWN | Negative | | |
| S mansoni 200pg | A caninum | UNKNOWN | Negative | | |
| S mansoni 200pg | A caninum | UNKNOWN | Negative | | |
| S mansoni 200pg | A caninum | UNKNOWN | Negative | | |
| Mixed Microb 200pg | A caninum | UNKNOWN | Negative | | |
| Mixed Microb 200pg | A caninum | UNKNOWN | Negative | | |
| Mixed Microb 200pg | A caninum | UNKNOWN | Negative | | |
| Human 200pg | A caninum | UNKNOWN | Negative | | |
| Human 200pg | A caninum | UNKNOWN | Negative | | |
| Human 200pg | A caninum | UNKNOWN | Negative | | |
| E coli 200pg | A caninum | UNKNOWN | Negative | | |
| E coli 200pg | A caninum | UNKNOWN | Negative | | |
| E coli 200pg | A caninum | UNKNOWN | Negative | | |
| Acan+ 20pg | A caninum | UNKNOWN | 19.178 | 19.143 | 0.049 |
| Acan+ 20pg | A caninum | UNKNOWN | 19.087 | 19.143 | 0.049 |
| Acan+ 20pg | A caninum | UNKNOWN | 19.163 | 19.143 | 0.049 |
| Acan+ 2pg | A caninum | UNKNOWN | 22.115 | 22.201 | 0.150 |
| Acan+ 2pg | A caninum | UNKNOWN | 22.373 | 22.201 | 0.150 |
| Acan+ 2pg | A caninum | UNKNOWN | 22.114 | 22.201 | 0.150 |
| Acan+ 200fg | A caninum | UNKNOWN | 26.142 | 25.854 | 0.338 |
| Acan+ 200fg | A caninum | UNKNOWN | 25.481 | 25.854 | 0.338 |
| Acan+ 200fg | A caninum | UNKNOWN | 25.939 | 25.854 | 0.338 |
| NTC | A caninum | NTC | Negative | | |
| NTC | A caninum | NTC | Negative | | |
| NTC | A caninum | NTC | Negative | | |

### DNA Extraction Protocol.

**Standard Operating Procedure for Processing Environmental Soil Samples (Modified from the Qiagen DNeasy PowerMax Soil DNA extraction protocol and the Sigma Millipore Pellet Paint NF DNA Concentration and Precipitation Protocol).**

**Procedure Overview:** This SOP is intended to facilitate the processing of environmental soil samples for the purpose of extracting soil transmitted helminth (STH) DNA.

**Required Equipment and Reagents (not included with the PowerMax Soil Kit):**

- 10% Bleach solution
- Wash bottle for bleach solution
- Latex or nitrile gloves
- Sterile 10 mL serological pipettes
- Pipette controller or bulb
- Disposable spatulas (Chemglass Life Science s, cat. no. CG-1985-P-07)
- Micropipettors and filter tips; p10, p20, p200 and p1000
- Sterile 50 mL conical-bottom centrifuge tubes
- Centrifuge capable of spinning 50 mL tubes at a minimum of 2,500 x g
- Vortex mixer
- 6 tube vortex adapter (Scientific Industries, cat. no. SI-H506)
- Waste beakers or other suitable liquid waste collection vessels
- Autoclavable hazardous waste collection bags
- Ethanol, absolute (100%), RNA/DNA grade
- 3M Sodium Acetate, pH 5.2
- PelletPaint NF Co-precipitant (Millipore Sigma cat. no. 70748-3)
- pDMD801 control
- Nuclease-free water
- Microcentrifuge tube racks
- Permanent markers

**The Qiagen PowerMax Soil Kit includes the following (designed for 10 extractions):**

- 10 MB Maxi Spin Columns
- 10 PowerMax Bead tubes
- 200mL PowerBead solution
- 2 x 6.6 mL Solution C1
- 2 x 28 mL Solution C2
- 44 mL Solution C3
- 330 mL Solution C4
- 4 x 30 mL Solution C5
- 66 mL Solution C6
- 40 x 50 mL Collection Tubes

**Note: All work is to take place in a Bio-Safety cabinet (BSC).** Be sure to wear gloves throughout the sampling procedure and change them frequently to avoid contamination between samples. Shake to mix Solution C4 before use. If Solution C1 has precipitated, heat at 60°C until precipitate dissolves.

**Note: An extraction blank (reagent only extraction) should be periodically performed (approximately once per week).** Performance of an extraction blank controls for cross-contamination of samples during the extraction procedure.

**Procedure:**

1. Use freshly-made 10% bleach to clean all equipment before bringing it into the BSC for use.
2. UV the BSC prior to starting the procedure. If a BSC is unavailable, designate an isolated bench space for dedicated use for soil extractions. Thoroughly bleach all surfaces within this dedicated space using freshly-made 10% bleach.
3. Add 15 mL of PowerBead Solution to each PowerMax Bead Tube.
4. Utilizing a disposable spatula, add 20 g of soil sample to each PowerMax Bead Tube containing PowerBead Solution.
5. Vortex at top speed for 1 minute.
6. Add 1.2 mL of Solution C1 to each PowerMax Bead Tube and vortex for 30 seconds.
7. Place each PowerMax Bead Tube on a vortex adapter (Scientific Industries cat. no. SI-H506) and vortex for 30 minutes at top speed.
8. Centrifuge at 2500 x g for 3 minutes at room temperature.
9. Divide the supernatant into two equal parts by pouring the supernatant evenly into two clean 50mL Collection Tubes (provided) leaving the pellet behind.
10. Add 2.5 mL of Solution C2 to each tube. Invert twice to mix.
11. Carefully pour the sample from one 50 mL sample tube into a new, clean 15 mL sample tube.
12. Label the 15 mL tube so that the sample ID matches that of the corresponding sample in the 50 mL tube (For later QA/QC purposes, it is critical that the sample IDs match).
13. Archive the sample in a 15 mL tube at -20°C for later processing (QA/QC, re-extraction, etc.). Proceed to step #14 with the remaining sample tube.
14. Incubate at 2–8°C for 10 minutes.
15. Centrifuge at 2500 x g for 4 minutes at room temperature.
16. Avoiding the pellet, pour the supernatant into a clean 50 mL Collection Tube (provided).
17. Add 4 mL of Solution C3 to each tube and invert twice to mix.
18. Incubate at 2–8°C for 10 minutes.
19. Centrifuge the tubes at 2500 x g for 4 minutes at room temperature.
20. Avoiding the pellet, pour supernatant into a clean 50 mL Collection Tube (provided).
21. Shake to mix Solution C4.
22. Add 30 mL of Solution C4 to each sample's supernatant and invert twice.
23. Add 1.0 µl of internal control (pDMD801 plasmid, 100pg / µl) to each sample.
24. Fill an MB Maxi Spin Column with the resulting solution from Step 23.
25. Centrifuge at 2500 x g for 2 minutes at room temperature.
26. Discard the flow-through into a waste container and add a second volume of supernatant to the same MB Maxi Spin Column.
27. Centrifuge again at 2500 x g for 2 minutes at room temperature.
28. Discard the flow-through into a waste container. ***Repeat steps 24-28 until the entire volume has been processed. This may require up to 4 total spins.***
29. Add 10 mL of Solution C5 to the MB Maxi Spin Column.
30. Centrifuge at 2500 x g for 3 minutes at room temperature.
31. Discard the flow-through into a waste container.
32. Centrifuge the column again at 2500 x g for 5 minutes at room temperature to fully dry the column.

33. Carefully place the MB Maxi Spin Column into a new 50mL Collection Tube (provided). Avoid splashing Solution C5 onto the column.
34. Add 5 mL of sterile Solution C6 to the center of the MB Maxi Spin Column membrane and centrifuge at 2500 x g for 3 minutes at room temperature.
35. Repeat Step 34 by adding the eluent back onto the column and centrifuge at 2500 x g for an additional 3 minutes at room temperature.
36. Discard the MB Maxi Spin Column. Extracts can be stored at -20 degrees C at this point or proceed with the following concentration and precipitation steps.
37. Thaw Pellet Paint NF Co-Precipitant (Sigma Millipore cat. no. 70748-3) and 3 M Sodium Acetate, pH 5.2, and bring to room temperature.
38. Add 5 µl of Pellet Paint NF Co-Precipitant to each sample, followed by 500 µL of 3 M sodium acetate, pH 5.2.
39. Add 10 mL of 100% ethanol to each sample.
40. Vortex briefly.
41. Incubate at room temperature for 2 minutes.
42. Centrifuge at top speed for 5 minutes.
43. A dark blue pellet should be visible at the bottom of tube. Without disturbing the pellet, carefully decant as much of the supernatant as possible into a waste beaker.
44. Using a serological pipet wash the pellet with 10 mL of 70% ethanol.
45. Vortex briefly.
46. Centrifuge at top speed for 5 minutes.
47. Remove the supernatant.
48. Using a serological pipet, wash the pellet with 10 mL of 100% ethanol.
49. Vortex briefly.
50. Centrifuge at top speed for 5 minutes.
51. Remove the supernatant.
52. With the lid open, air dry the pellet overnight in an isolated location (preferably a hood).
53. Add 200 µL of nuclease-free water to the isolated/dry pellet. Using a clean P200 pipet tip for each sample, gently pipet up and down to aid resuspension.
54. Allow the pellet to resuspend for a minimum of 4 hours at room temperature
55. Following the 4 hour resuspension, using a clean P200 pipet tip for each sample, gently pipet up and down again to ensure resuspension is complete.
56. Transfer the entire 200 µL sample to a new, clean cryovial.
57. Label the cryovial with the same sample number as the parent tube.
58. Store the samples at -80 °C if available. If unavailable, -20 °C storage is acceptable.

### qPCR Protocol.

**Standard Operating Procedure for the Real-Time PCR analysis of Samples for the Presence of Soil-Transmitted Helminths**

**Procedure Overview**: This procedure is used for DNA amplification of specific targets using primer/probe for the diagnosis of soil-transmitted helminths (STH).

**Equipment and reagents necessary**:
- Micropipettors and tips (p1000, p200, p20, p10)
- 96-well Semi-skirt 0.1 mL PCR Plates (USA Scientific # 1402-9100)
- Ice buckets, ice
- 1.7 mL microfuge tubes for master mix and NTC (Phenix Research Products # MAX-715)
- Adhesive optical film for qPCR (USA Scientific # 2921-7800)
- Temporary adhesive film for transport (USA Scientific # 2920-3500)
- Adhesive film applicator (ThermoFisher Scientific # 4333183)
- Permanent marker
- Microfuge tube openers (USA Scientific # 1400-1508)
- 10 µM stocks of Forward/Reverse primers and probes (see recipes for master mix below)
- Real-time PCR thermocycler (We recommend Applied Biosystem's StepOnePlus Real-time PCR System)
- Molecular biology grade water
- 0.1X Tris-EDTA solution
- TaqPath ProAmp PCR Master Mix (ThermoFisher Scientific # A30867)
- DNA plasmid templates (stock at 10 pg/µL) - obtained from the Williams Lab
- Aluminum foil
- Nitrile or latex gloves
- Standard nanofuge
- Microcentrifuge tube racks

**Note:** Wear gloves and change them FREQUENTLY to avoid contamination between samples; be certain that master mix preparation occurs in an isolated master mix preparation area (dead air box) and that no other work occurs in this location.

**Pre-Procedure Steps:**
1. Prepare aliquots of TaqPath ProAmp Master Mix in accordance with **Appendix A.**
2. Prepare primer working stocks in accordance with **Appendix B**.

**Procedure:**
1. Using 10% bleach (make this freshly each day) disinfect your tip boxes, pipettes, ice buckets, pens/markers, racks, bags of tubes, and anything that you might be using in the pre-PCR station dead air box.
2. **UV** sterilize the bleached dead air box containing your bleached set of pipettes/tips, racks, markers, etc. for **20 minutes.**
3. Following the completion of the UV cycle, place your 350 µL aliquot of TaqPath Pro Amp Master Mix, your primer and probe working stocks (10 µM) and aliquot of PCR grade/nuclease free water on ice. Place your ice bucket in the dead air box and allow all reagents to thaw completely (approximately 20 min).

4. Prepare your template addition station by bleaching your pipets, tips, racks, and the working station itself. UV all instruments, etc. that will be used during template addition, as well as the template addition station dead air box itself.
5. Turn on the StepOnePlus instrument to warm up.
6. Following the completion of the UV cycle at the template addition station, thaw your samples and your aliquot of positive control plasmid in the pre-PCR station dead air box. (As samples are DNA, it is not necessary to thaw them on ice.)
7. Following the completion of the 20 min UV sterilization cycle in the pre-PCR room, use micropipettors to prepare a pooled reaction mix for 100 reactions (see table below for recipes per assay per 100 reactions).
8. Aliquot 5 µL of prepared reaction mix into each well of the 96-well plate.
9. Rest a temporary adhesion film on top of your plate (to protect the wells from exposure to particulates in the air) and transfer your plate to the template addition area. **Leave all pre-PCR equipment and reagents in the pre-PCR station dead air box. DO NOT take any of these materials with you to the template addition area.**
10. In the template addition dead air box, prepare your plasmid control dilutions in accordance with **Appendix C**.
11. After all sample tubes are thawed, flick all tubes to mix and briefly spin all tubes in a nanofuge to ensure that each tube's sample volume is not near the cap of the tube.
12. Add 2 µL of DNA template (unknown sample/reaction blank/plasmid control/NTC control) to each well.
    a. **Follow the "unified plate layout found in Table 2" when adding template.**
    b. **Use a new, clean/bleached tube opener to open each sample tube and immediately discard the tube opener into the soiled opener receptacle.**
13. Following the addition of all samples and controls, apply an optical reaction plate cover to each plate and carefully seal the plate using the plate sealer. To prevent evaporation during the real-time PCR run, be certain to fully seal the plate along all of its edges.
14. Spin down the 96-well plate on a clinical centrifuge (~2,000 rpm) for 1 minute. Be sure the centrifuge is perfectly balanced. **(Note:** If a clinical centrifuge is not available, tap the plate on your bench when finished loading the template and sealing the plate with the film to pop any bubbles seen within the wells.)
15. Set up the PCR program on the computer (**Note: This description is for use with the Applied Biosystems StepOnePlus Instrument. Steps #15-17 will vary depending upon the specific instrument used**):
    - **File → New Experiment → Advanced Setup**
    - **Experimental properties:**
        - *Instrument*: StepOnePlus Instrument (96 wells)
        - *Type of experiment*: Quantitation – Comparative Ct (ΔΔCt)
        - *Reagents*: TaqMan Reagents
        - *Ramp speed*: Standard (~ 2 hours to complete a run)
    - **Plate set-up:**
        - *Define targets and samples*:
            - Name your target
            - Define your reporter/quencher for the probe (For all of the assays you should select FAM-None as the Dye-Quencher combination.)
            - Create your samples and name them
        - *Assign targets to wells*: match each well with the appropriate sample name defined in the previous step. Reassign your "No Template Control" (NTC) wells to be "Negative" rather than "Unknown" samples.
    - **Run method:**

- *Volume of reaction:* For all the STH assays, you should select 7 µl reaction as the reaction volume.
- *Cycling conditions:* For all the STH assays, 59ºC should be selected instead of 60ºC (the default) as the annealing and extension temperature for 1 minute.
  - Initial 2 min incubation step at 50°C, followed by a 10 min incubation at 95°C
  - 40 cycles of:
    - 95°C for 15 sec for denaturation
    - 59°C for 1 min for annealing and extension

- **Save your run file:**
  - Use the following naming convention when saving all run files:
    - Studyname_Target_Firstsample_Lastsample_Date_Technician Initials
      - Ex – Ethiopia_Ad_2-1_2-42_June-5-2018_JAM
- **<u>Note:</u>** To save time, plate set-up information can be saved in advance as a template and/or imported directly from a Microsoft Excel file.
  - Alternatively, a run "template" can be created. To create a run template, follow the PCR program set-up instructions above, but save your file as a "template". After saving, close the template file, then re-open it to begin your run. To create a file for each subsequent run, re-open the template file and simply redefine your sample list such that the sample order in the new list matches the sample order from the template file. For example, the sample which appears in wells A1 and A2 in the template file should be replaced by the sample which you desire to appear in wells A1 and A2 in the new file. The redefined samples will then auto-populate into the template's plate map, replacing the original samples. Save your new template with a unique name using the naming convention described above.

16. Run your plate on the real-time PCR instrument.
17. Once the run has completed, perform the following data collection/analysis steps:
    - Manually set the threshold so that it crosses any amplification curves during the "early exponential" phase of amplification.
    - Click the green "analyze" button.
    - Save the completed run using the same naming convention described above, with the addition of "_POST" to the end of the file name. For example: "Ethiopia_Ad_2-1_2-42_June-5-2018_JAM_POST".
    - Export results by selecting File → Export → Results. Then click the "Browse" button to select the location to export your results to. Once the location is selected, click "Open". You may then save your data using the default file name. (Note: this name will be the same as your pre-run file name, with the "_data" designation automatically added at the end upon saving).

**Table 1. Recipe for Reaction Mixes for Each STH Assay (100 Reactions)**

| | *N. americanus* | *T. trichiura* | *A. lumbricoides* | *A. duodenale* | *A. ceylanicum* | **Control Plasmid** |
|---|---|---|---|---|---|---|
| **Final Primer Concentrations in Reaction Mix** | F/R 250 nM | F 62.5 nM / R 250 nM | F/R 62.5 nM | F/R 250 nM | F 125 nM / R 1000 nM | F/R 250 nM |
| **TaqPath ProAmp Master Mix** | 350 µL (use P1000) | 350 µL (use P1000) | 350 µL (use P1000) | 350 µL (use P1000) | 350 µL (use P1000) | 350 µL (use P1000) |
| **$ddH_2O$** | 136.3 µL (use P200 for 130 µL and P10 for 6.3 µL) | 149.4 µL (use P200 for 140 µL and P10 for 9.4 µL) | 127.55 µL (use P200 for 120 µL and P10 for 7.55 µL) | 136.3 µL (use P200 for 130 µL and P10 for 6.3 µL) | 92.55 µL (use P200 for 90 µL and P10 for 2.55 µL) | 136.3 µL (use P200 for 130 µL and P10 for 6.3 µL) |
| **Forward primer** | 17.5 µL (use P20) | 4.4 µL (use P10) | 8.75 µL (use P10) | 17.5 µL (use P20) | 8.75 µL (use P10) | 17.5 µL (use P20) |
| **Reverse primer** | 17.5 µL (use P20) | 17.5 µL (use P20) | 35 µL (use P200) | 17.5 µL (use P20) | 70 µL (use P200) | 17.5 µL (use P20) |
| **Probe (125 nM)** | 8.7 µL (use P10) | 8.7 µL (use P10) | 8.7 µL (use P10) | 8.7 µL (use P10) | 8.7 µL (use P10) | 8.7 µL (use P10) |

**Table 2. Unified Plate Layout:**

| | 1 | 2 | 3 | 4 | 5 | 6 | 7 | 8 | 9 | 10 | 11 | 12 |
|---|---|---|---|---|---|---|---|---|---|---|---|---|
| A | 1 | 1 | 2 | 2 | 3 | 3 | 4 | 4 | 5 | 5 | 6 | 6 |
| B | 7 | 7 | 8 | 8 | 9 | 9 | 10 | 10 | 11 | 11 | 12 | 12 |
| C | 13 | 13 | 14 | 14 | 15 | 15 | 16 | 16 | 17 | 17 | 18 | 18 |
| D | 19 | 19 | 20 | 20 | 21 | 21 | 22 | 22 | 23 | 23 | 24 | 24 |
| E | 25 | 25 | 26 | 26 | 27 | 27 | 28 | 28 | 29 | 29 | 30 | 30 |
| F | 31 | 31 | 32 | 32 | 33 | 33 | 34 | 34 | 35 | 35 | 36 | 36 |
| G | 37 | 37 | 38 | 38 | 39 | 39 | 40 | 40 | 41 | 41 | 42 | 42 |
| H | 10 pg/ µL cont | 10 pg/ µL cont | 100 fg/ µL cont | 100 fg/ µL cont | 1 fg/ µL cont | 1 fg/ µL cont | NTC | NTC | NTC | NTC | Repeat or extr blank control | Repeat or extr blank control |

Grant, Jessica R., Nils Pilotte, and Steven A. Williams. 2019. "A Case for Using Genomics and a Bioinformatics Pipeline to Develop Sensitive and Species-Specific PCR-Based Diagnostics for Soil-Transmitted Helminths." *Frontiers in Genetics* 10 (September): 883.

Pilotte, Nils, Marina Papaiakovou, Jessica R. Grant, Lou Ann Bierwert, Stacey Llewellyn, James S. McCarthy, and Steven A. Williams. 2016. "Improved PCR-Based Detection of Soil Transmitted Helminth Infections Using a next-Generation Sequencing Approach to Assay Design." *PLoS Neglected Tropical Diseases* 10 (3): e0004578.
